# Genome-wide analysis identifies 66 variants underlying anatomical variation in human neuroendocrine structures and reveals links to testosterone

**DOI:** 10.1101/2024.08.01.24311295

**Authors:** Hannah Currant, Christopher Arthofer, Teresa Ferreira, Gwenaelle Douaud, Barney Hill, Samvida S Venkatesh, Nikolas A Baya, Duncan S Palmer, Saskia Reibe, Anje Moltke-Prehn, Tune H Pers, Andreas Bartsch, Jesper Andersson, Margaret F Lippincott, Yee-Ming Chan, Stephanie B Seminara, Thomas E Nichols, Christoffer Nellaker, Stephen Smith, Søren Brunak, Frederik J Lange, Cecilia M Lindgren

**Affiliations:** Nuffield Department for Population Health, University of Oxford, Old Road Campus, Oxford, OX3 7LF, UK; Big Data Institute, University of Oxford, Old Road Campus, Oxford, OX3 7LF, UK; Novo Nordisk Foundation Center for Protein Research, University of Copenhagen, Blegdamsvej, Copenhagen, 2200, Denmark; Oxford Centre for Functional MRI of the Brain, University of Oxford, John Radcliffe Hospital, Oxford, OX3 9DU, UK; Wellcome Centre for Human Genetics, University of Oxford, Old Road Campus, Oxford, OX3 7LF, UK; Novo Nordisk Foundation Center for Basic Metabolic Research, University of Copenhagen, Blegdamsvej, Copenhagen, 2200, Denmark; Department of Neuroradiology, University of Heidelberg, Im Neuenheimer Feld, Heidelberg, 69120, Germany; Reproductive Endocrine Unit, Massachusetts General Hospital, Fruit Street Gray, Boston, 02114, MA, USA; Harvard Center for Reproductive Medicine, Massachusetts General Hospital, Fruit Street Gray, Boston, 02114, MA, USA; Division of Endocrinology, Boston’s Children’s Hospital, Longwood Avenue, Boston, 02115, MA, USA

**Author notes:** Contributing authors. These authors contributed equally to this work (Hannah Currant, Christopher Arthofer, Frederik J Lange, Cecilia M Lindgren).

**Keywords:** endocrinology, brain anatomy, image-derived phenotypes, genetic association studies

## Abstract

The hypothalamus, pituitary gland and olfactory bulbs are neuroanatomical structures key to the regulation of the endocrine system. Variation in their anatomy can affect the function of the reproductive system. To investigate this relationship, we extracted four largely unexplored phenotypes from 34,834 individuals within UK Biobank by quantifying the volume of the hypothalamus, pituitary gland and olfactory bulbs using multi-modal magnetic resonance imaging. Genome-wide association studies of these phenotypes identified 66 independent common genetic associations with endocrine-related neuroanatomical volumes (***P <* 5 *×* 10*^−^*^8^**), five of which had a prior association to testosterone levels, representing enrichment of testosterone-associated SNPs over random chance (***P*** -value = **9.89 *×* 10*^−^*^12^**). Exome-wide rare variant burden analysis identified *STAB1* as being significantly associated with hypothalamus volume ( ***P* = 3.78 *×* 10*^−^*^7^**), with known associations to brain iron levels. Common variants associated with hypothalamic grey matter volume were also found to be associated with iron metabolism, in which testosterone plays a key role. These results provide initial evidence of common and rare genetic effects on both anatomical variation in neuroendocrine structures and their function in hormone production and regulation. Variants associated with pituitary gland volume were enriched for gene expression specific to theca cells, responsible for testosterone production in ovaries, suggesting shared underlying genetic variation affecting both neuroanatomical and gonadal endocrine tissues. Cell-type expression enrichment analysis across hypothalamic cell types identified tanycytes to be associated (***P* = 1.69 *×* 10*^−^*^3^**) with olfactory bulb volume associated genetic variants, a cell type involved in release of gonadotropin-releasing hormone into the bloodstream. Voxel-wise analysis highlighted associations between the variants associated with pituitary gland volume and areas of intracranial venous drainage involved in hormonal release into the blood circulation. Together, our results suggest a shared role of genetics impacting both the anatomy and function of neuroendocrine structures within the reproductive system in their production and release of reproductive hormones.

## Introduction

The endocrine system is integral to reproductive function, with its disruption impacting development, fertility and broader disease. The intricate task of regulating the endocrine system is orchestrated by neuroanatomical structures. Three neuroanatomical structures are of key importance: (1) the hypothalamus, which contains specialised neurons that produce gonadotropin releasing hormone (GnRH); (2) the pituitary gland, which responds to GnRH and aids regulation of hormone production [1]; and (3) the olfactory bulbs, along which the GnRH releasing neurons migrate during fetal development [2]. The pituitary gland, when stimulated by GnRH, produces luteinising hormone (LH) and follicle stimulating hormone (FSH) which, in turn, regulate the production of other sex hormones such as testosterone, progesterone and oestrogen [3]. Collectively, these hormones control reproductive processes including puberty, fertility, menstruation and menopause.

Variation in the anatomy (specifically, the size/volume) of these three neuroanatomical structures occurs within the population and has been reported to be associated with variation in their function within the reproductive system across a spectrum of severity. For example, population-level common variation in the volume of the pituitary gland has been found to be correlated with sex-steroid concentration [4]. At the more pathological end of the spectrum, individuals with Kallmann syndrome exhibit hypoplasic or absent olfactory bulbs, alongside additional characteristics including delayed or absent onset of puberty, infertility and anosmia [5].

Magnetic resonance imaging (MRI) offers the opportunity to accurately characterise anatomical variation in brain structure. Further, biobanks containing such imaging data allow us to do so at a unprecedented scale. Novel and advancing methodologies enable quantification of anatomical variation at such scale, offering opportunities for well-powered statistical analyses of subtle differences [6]. Image-derived phenotypes (IDPs), such as the volumes of anatomical structures, have proven a valuable resource for enabling genetic discovery across numerous biological systems including the cardiac [7], ophthalmic [8], and neurological [6] systems. By extracting low-dimensional phenotypes from high-dimensional data (MRI images), based on *a priori*knowledge of bio-logical relationships, there is the opportunity to increase statistical discovery power in genetic association studies.

Examining the genetic contributors to anatomical variation in these neuroendocrine structures provides an opportunity to further our understanding of their underlying biology and their function. In doing so, we aim to elucidate their connection to the broader endocrine and reproductive systems. To do this, we extracted quantitative IDPs measuring structure volume from MRI images of 34,834 individuals within UK Biobank (UKB), representative of the European subset (Methods) of individuals with MRI available. We performed common-variant genome-and rare-variant exome-wide association studies of these neuroendocrine IDPs. This is, to our knowledge, the first population-scale study of IDPs relating to these neuroendocrine structures and the largest study of genetic effect on their anatomy by far. We hypothesise that use of these IDPs at unprecedented scale will allow for discovery of novel genetic variants affecting neuroanatomy, and additional understanding of links between the neurological and endocrine systems through common biological pathways.

## Results

### Extraction of phenotypes describing neuroendocrine anatomy

The volumes of the hypothalamus, pituitary gland and olfactory bulbs (combined left and right) were extracted with a label propagation approach from the MRI images of 34,834 individuals following quality control (Fig. 1A). Hypothalamic grey matter volume was extracted directly from grey matter probability maps (see Methods for details of image-derived phenotype extraction). All measurements were taken as absolute volumes (without normalisation for brain size) while accounting for partial volumes after label propagation (see Methods 10). The hypothalamus is the largest of the four volumes (mean = 466.16mm^3^ *±* 50.84 (SD)), followed by the hypothalamic grey matter (mean = 456.31mm^3^ *±*62.06), pituitary gland (mean = 268.02mm^3^ *±*64.15) and olfactory bulbs (mean = 62.68mm^3^ *±* 17.55). Across all four IDPs, males had higher volumes than females, in keeping with prior knowledge [9] (Fig. A1, Table A1). We adjusted the IDPs for a number of variables including age, sex and head size (for complete details see genome-wide association studies Methods) to carry forward for further analysis.

**Fig. 1.**
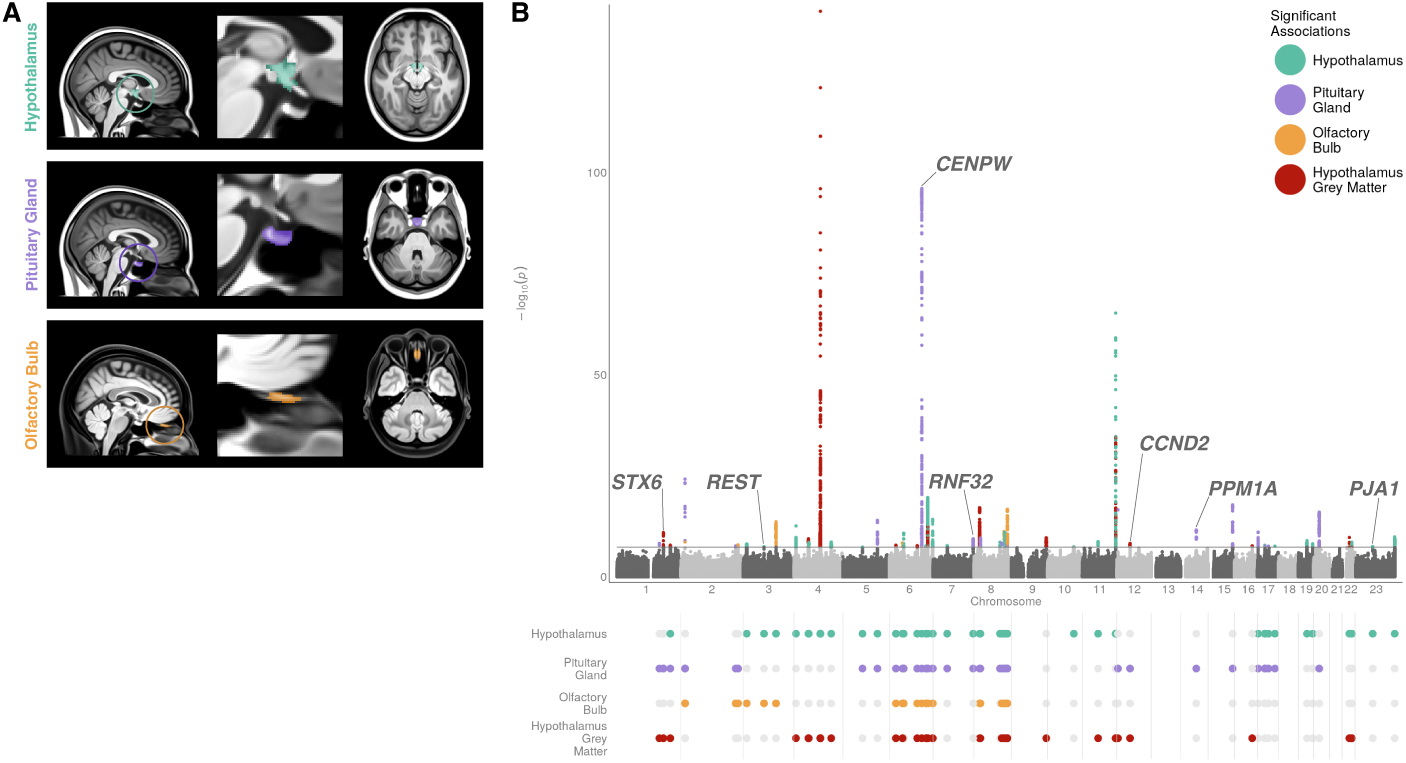
GWAS of the volume of four neuroendocrine structures (hypothalamus, pituitary gland, olfactory bulb, and hypothalamic grey matter volume) extracted from MRI images. A) Atlas consisting of a standard brain template with the endocrine-associated neuroanatomical segmentations overlaid in colour, as circled in the left of the plot. B) The upper panel is a Manhattan plot presenting the result of four GWAS of hypothalamus (turquoise), pituitary gland (purple), mean of left and right olfactory bulbs (gold), and hypothalamic grey matter (red) volume, in a sex-combined sample. Each point represents a genetic variant with the *x*-axis representing the genomic position within each chromosome and the *y*-axis representing the *−* log_10_(*P* -value) of the association in an additive linear model. Variants with a prior association to testosterone or menstrual timing are highlighted and their gene name listed. The lower panel is an upset plot representing the overlap of the signals across the GWAS of the four neuroendocrine traits. The *x*-axis represents the genomic position, aligned with the Manhattan plot. The presence of an overlap between the signal is shown with a coloured dot, no overlap is shown with a grey dot.

### Genetic variation associated with anatomical variation in neuroendocrine structures

We conducted the largest to-date genome-wide association studies (GWAS) of four under-explored IDPs describing the volume of the neuroendocrine struc-tures across ten million SNPs (minor allele frequency (MAF) *>*0.01). We identified 66 independent lead variants significantly associated (*P <* 5 *×* 10*^−^*^8^) with one of the four neuroendocrine IDPs in a sex-combined sample: 26 asso-ciated with hypothalamus volume, 16 with pituitary gland volume, 6 with olfactory bulb volume, and 18 with hypothalamic grey matter volume (Fig. 1B, Table A2). Application of LD-score regression (LDSC) [10] showed all GWAS were controlled for residual population structure (Fig. A2, Table A4, maximum LDSC intercept = 1.02, minimum LDSC intercept *P* = 0.02).

In five cases, using conditional joint analysis using GCTA-COJO [11], we identified two or more independent signals within the same locus. Although lead SNPs were unique to each phenotype, conditional analysis across GWAS identified a number of loci with overlapping genetic association signal across the four IDPs (Fig. 1B). Genome-wide SNP-based heritability (*h*^2^ *±* stan-dard error (SE)), was calculated [10] for hypothalamus (0.25 *±* 0.02), pituitary gland (0.20 *±* 0.03), olfactory bulb (0.11 *±* 0.02), and hypothalamic grey matter volume (0.25 *±* 0.02).

### Sexual dimorphism in genetic effect

We observed sexual dimorphism in sex-stratified GWAS (18,487 females and 16,347 males, Fig. A3): among lead variants significantly associated (*P <* 5 *×* 10*^−^*^8^) with each of the four neuroendocrine IDPs in the sex-combined, female or male populations, four variants displayed a statistically significant difference (*P <* 5.5 *×* 10*^−^*^4^) in effect size between females and males (Fig. A4A, Table A5). These were: rs6544040, in an intronic region of *VIT* (hypothalamus volume females: *β* = 0.07 and SE = 0.01, males: *β* = *−*0.002 and SE = 0.01; sex difference *P* = 1.31 *×* 10*^−^*^5^); rs186990314, an intronic variant in *SMR3B* (pituitary gland volume females: *β* = 0.001 and SE = 0.02, males: *β* = *−*0.09 and SE = 0.02; sex difference *P* = 4.22 *×* 10*^−^*^5^); rs7749444, an intronic variant near *PRIM2BP* (olfactory bulb volume females: *β* = 0.01 and SE = 0.01, males: *β* = 0.06 and SE = 0.01; sex difference *P* = 1.17 *×* 10*^−^*^3^); and rs144968764, an intronic variant near *MND1* (hypothalamic grey matter volume females: *β* = *−*0.24 and SE = 0.04, males: *β* = 0.01 and SE = 0.05; sex difference *P* = 6.65 *×* 10*^−^*^5^). Common genetic variants in *VIT* have been associated with cortical surface area asymmetry [12]. Additionally, comparison of the SNP-based heritability [10] of each of the traits across the sexes showed a nominally significant difference in the SNP-heritability of olfactory bulb volume (*t*-test *P* = 0.04, Fig. A4B), with females having a higher heritability (*h*_SNP_^2^ = 0.16 *±* 0.03) than males (*h*_SNP_^2^ = 0.07 *±* 0.03).

### Rare variant association testing across the exome

To assess the rare variant contribution to neuroendocrine volumes, we carried out rare-variant association studies (RVAS) across the exome at the variant-(N=32,537,048) and gene-level (N=19,489) for each of the four neuroendocrine IDPs. Following multiple testing correction (*P <* 6.25 *×* 10*^−^*^7^), rare damaging variation in a single gene, *STAB1*, was found to be associated (Cauchy combination test [13] of all damaging variants across all MAF thresholds, *P* = 3.78 *×* 10*^−^*^7^) with hypothalamus volume (Fig. 2, Methods). Within STAB1 we found a particularly strong association (P=7.73 *×* 10*^−^*^8^) subset-ting to ultra-rare (*MAF ≤* 1 *×* 10*^−^*^4^) pLoF or damaging missense variants with hypothalamus volume. No common variants (MAF *>*0.01) within this gene were significantly associated with hypothalamus volume in our GWAS. GWAS have previously found variants mapped to this gene to be associated with vertex-wise sulcal depth [14] and cortical thickness [15]. *STAB1* encodes stabilin 1, a type 1 transmembrane receptor [16] which has been associated with brain iron levels [17] and functional brain measurements [18]. Brain iron levels and homeostasis are known to play a role in several neurological diseases including Parkinson’s [19], for which *STAB1* is also a gene of interest [20].

**Fig. 2.**
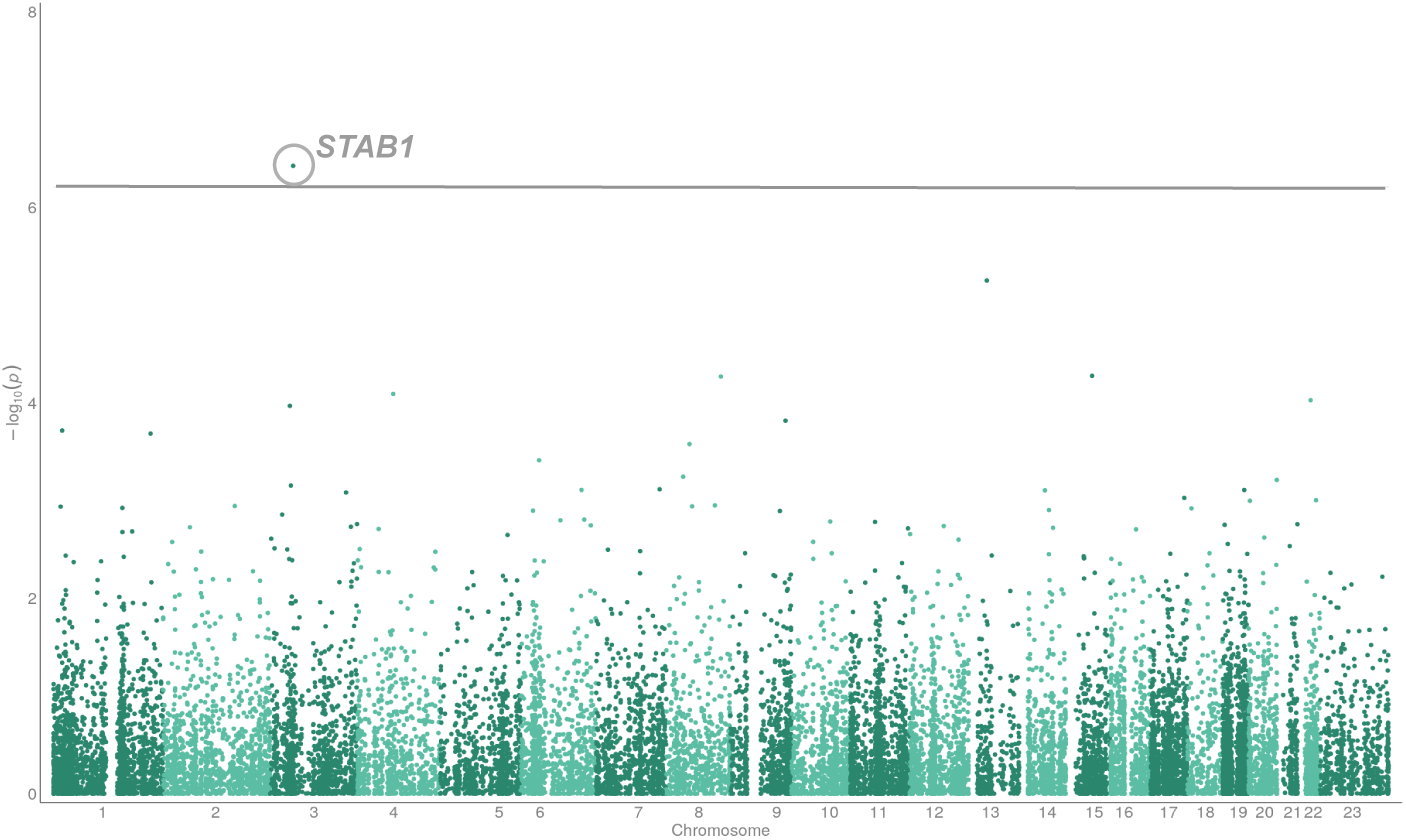
RVAS of hypothalamus volume. A Manhattan plot of RVAS of hypothalamus volume in a sex-combined sample using a cauchy combination test using damaging gene burden tests. Variants in odd and even chromosomes are coloured in dark and light green. The *y*-axis is the *−* log_10_(*P* -values) of association. The exome-wide significant (*P <* 6.25 *×* 10*^−^*^7^) gene, *STAB1* association is highlighted.

### Phenome-wide association analyses across clinical phenotypes

To explore the overlap between the 66 lead variants identified in our GWAS and a range of clinical phenotypes, we carried out phenome-wide association studies (PheWAS) across 1839 phecodes in the UKB by mapping International Statistical Classification of Diseases and Related Health Problems 10th Revi-sion (ICD10) codes (Methods). We chose to use phecodes to group ICD10 codes as they are designed to capture clinically meaningful concepts for research and tend to comprise a collection of ICD10 codes, resulting in larger case counts and so increased power compared to two-digit ICD10 codes (Fig. A5, Tables A6, A7, A8, A9) [21]. Following Benjamini-Hochberg (BH) [22] false discovery rate (FDR) multiple test correction, three phenotypes were significantly asso-ciated (BH FDR *P <* 0.05) with hypothalamic grey matter associated SNPs (Fig. 3): disorders of iron metabolism (*P* = 3.27 *×* 10*^−^*^65^), disorders of min-eral metabolism (*P* = 8.99 *×* 10*^−^*^45^) and sicca syndrome (*P* = 3.48 *×* 10*^−^*^6^). As highlighted following rare-variant association testing, iron homeostasis is integral to brain function with its dysfunction seen in several neurological diseases. Additionally, diseases of iron overload in males are associated with reproductive dysfunction including hypogonadotropic hypogonadism and infertility [23]. Further, testosterone levels are known to be associated with iron levels, with testosterone playing a key role in erythropoiesis and thus iron incorporation and homeostasis [24]. Meanwhile, sicca syndrome is a condition affecting production of fluids including saliva and tears. It has previously been shown to have an association with oestrogen levels [25] and is most commonly diagnosed in postmenopausal women [26]. These associations could suggest a relationship between the structures producing reproductive hormones and their downstream effects across biological systems.

**Fig. 3.**
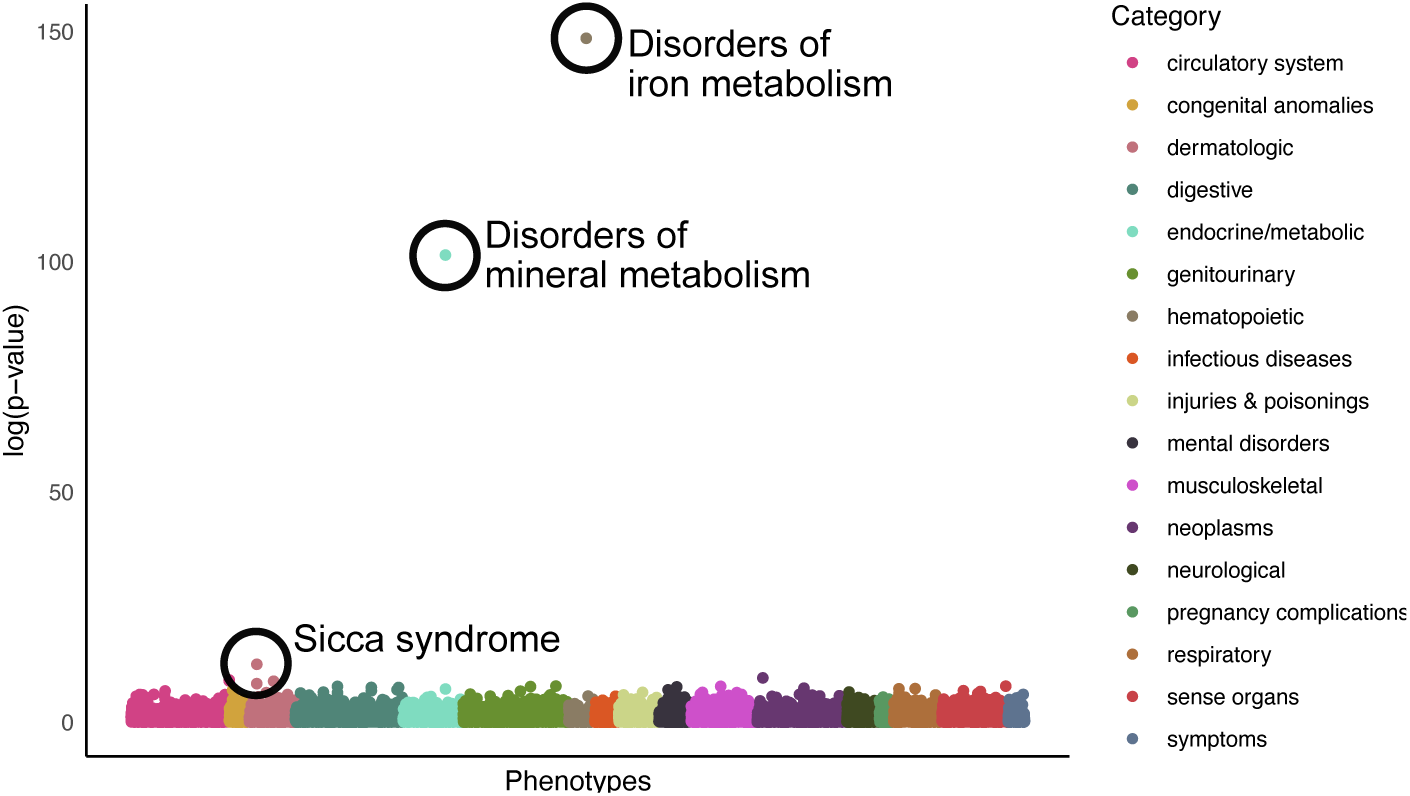
PheWAS of 18 significant GWAS associations. Scatter plot displaying the *−* log_10_(*P* -values) from a PheWAS of 1839 phecodes extracted from UKB (Methods) and the 18 significant lead variants identified as associated with the volume of the hypothalamic grey matter. Statistically significant associations following Benjamini-Hochberg multiple test correction (*P <* 0.05) are circled in black.

### Overlap of neuroendocrine genetics with neurological phenotypes

Ten of the 66 variants associated with the neuroendocrine IDPs have previously been found to be significantly (*P <* 5*×*10*^−^*^8^) associated with other brain phenotypes including substructure volumes, cortical thickness and brain connectivity (Table A3). To further explore the shared genetics underpinning brain structure phenotypes, we performed a PheWAS using Big40 [27], a database of GWAS summary statistics of *∼* 4, 000 multi-modal MRI-derived phenotypes within the UKB. This identified significant associations (*P <* 1.93 *×* 10*^−^*^7^, Bonferroni multiple testing correction applied accounting for 259,710 tests) with 823 of the neurological MRI IDPs (Fig. A6, Tables A10, A11, A12 & A13). Neuroendocrine IDP-associated genetic variants discovered in our study were associated with 226 regional and tissue volumes and intensities. The most significant associations were between rs13107325 (a missense variant in *SLC39A8*) and the volume of the left (*β* = 0.43, SE = 0.02, *P* = 2.09*×*10*^−^*^175^) and right ventral striatum (*β* = 0.41, SE = 0.02, *P* = 2.51 *×* 10*^−^*^165^), the volume of the left (*β* = 0.39, SE = 0.02, *P* = 8.91 *×* 10*^−^*^147^) and right (*β* = 0.40, SE = 0.02, *P* = 5.25 *×* 10*^−^*^155^) putamen, and the T1 intensity within the right hemisphere accumbens area (*β* = *−*0.40, *SE* = 0.02, *P* = 1.95 *×* 10*^−^*^146^). The overlap in associations of genetic variants with the neuroendocrine IDPs and broader anatomical brain phenotypes is to be expected due to shared developmental processes and common cell types, and has been seen between other brain phenotypes previously [6].

### Overlap of neuroendocrine genetics with testosterone and other reproductive phenotypes

#### Testosterone related genetic variants

Fourteen variants (21%) that we observed to be associated with the neuroendocrine IDPs have been previously reported to be associated with reproductive traits (Table A3). Notably, there was a significant enrichment within the variants associated with the IDPs for prior associations to testosterone (*P* -value = 9.89 *×* 10*^−^*^12^). Five loci have prior associations with testosterone: firstly, rs11941568 (hypothalamus volume *β* = 0.04, SE = 0.01, MAF = 0.40), an upstream variant of *REST* which is a transcriptional repressor that represses neuronal genes in non-neuronal tissues [28]. Secondly, a downstream intronic variant of *CENPW* (rs2184968, pituitary gland volume *β* = 0.16, SE =0.007, *MAF* = 0.45) which is also associated with male-pattern baldness [29], a phenotype known to be largely controlled by testosterone [30]. Thirdly, rs76895963 (pituitary gland volume *β* = 0.24, SE = 0.029, MAF = 0.02) in an intronic region of *CCND2* has a prior association with testosterone [31]. rs76895963 colocalised with an eQTL for *CCND2*, enriched in tissues including the pituitary gland, thyroid, tibial nerve and cerebellum [32]. *CCND2* deficient mice have hypoplasic testes in males and are sterile in females [33] with *CCND2* expression mirroring blood testosterone levels [34]. Further, it is thought that *CCND2* might help regulate spermatogenesis [34] and be involved in impotence [35]. Fourthly, a genetic variant upstream of *RNF32* (rs7808966, pituitary gland volume *β* = 0.03, SE = 0.01, MAF = 0.07), a gene expressed in both the ovaries and testis, and expressed during spermatogenesis [36]. Fifthly, rs10220706 (pituitary gland volume *β* = *−*0.06, SE = 0.01, MAF = 0.30), downstream of *PPM1A*, has prior associations to testosterone and also to age at menarche [29]. *PPM1A* has been shown to be involved in regulation of testosterone synthesis in Leydig cells within the testes [37].

### Reproductive development and ageing-related genetic variants

We also observed an overlap of variants found to be associated with the neuroendocrine IDPs with the genetics of menstrual timing: both age at menarche and age at menopause (Table A3). For example, a missense variant on chromosome X in *PJA1* (rs5937160, hypothalamus volume *β* = *−*0.04, SE = 0.01, MAF = 0.23) is associated with age at menarche in females, and the relative age of first facial hair and the relative age at which one’s voice broke in males [29]. Additionally, a locus located in *STX6*, associated with hypothalamus volume (rs1044595, *β* = *−*0.04, SE = 0.007, MAF = 0.40) and hypothalamic grey matter volume (rs35306826, *β* = *−*0.05, MAF = 0.41), has prior association to menopause status and the age at menopause, in addition to prior associations to other brain anatomy phenotypes [27, 38].

### Genetic relationships with reproductive traits

We evaluated genetic correlations (*r_g_*) between the neuroendocrine IDPs and reproductive hormone (testosterone, FSH, LH, oestradiol and progesterone [39]) levels (Fig. A14). We found nominally significant *r_g_* (*P <* 0.05) between testosterone level and pituitary gland volume in a sex-combined analysis (*P* = 0.03, *r_g_* = 0.07), and between testosterone levels and both hypothalamus (*P* = 9.9 *×* 10*^−^*^3^, *r_g_* = 0.11) and olfactory bulb volume (*P* = 0.01, *r_g_* = 0.13) in females. We also assessed *r_g_* between the neuroendocrine IDPs and six infertility phenotypes -five female infertility phenotypes (all cause, anatomical cause, anovulatory, unknown cause excluding idiopathic causes, and unknown cause including idiopathic fertility) and a male infertility phenotype [39]. None of the phenotype pairs displayed significant genetic correlation (*P <* 0.05).

We carried out bidirectional Mendelian randomisation analysis to investigate causality between the neuroendocrine IDPs, and the reproductive hormone and infertility phenotypes outlined above. Phenotypes with at least five genetic instruments were used as exposures. We found no evidence of a causal relationship between the neuroendocrine IDPs and any of the reproductive phenotypes following multiple test correction (*P <* 4.17 *×* 10*^−^*^4^ for all tests, Table A15).

We did not find any statistically significant evidence of colocalisation between genetic variants found to be associated with one of the neuroendocrine IDPs and reproductive hormones or fertility phenotypes (all 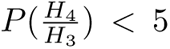 & P(*H*_4_) *<* 0.5).

### Tissue and pathway enrichment

The neuroendocrine structures studied here are involved in the production of reproductive hormones that act across many tissues. We therefore explored the colocalisation between the 66 neuroendocrine IDP genetic association signals with the tissue-specific expression quantitative trait loci (eQTLs) within the GTEx dataset [32]. We found that three genetic variants colocalised (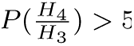 & P(*H*_4_) *>* 0.5) with eQTLs for genes known to be involved in sperm-oocyte fusion: rs4806665 with *TMEM190* enriched in the testes and lung [40], rs7550273 with *CD46* enriched in the oesophagus mucosa [41], and rs10800397 with *SPATA46* enriched in the brain cerebellum [42]. Six genetic variants also colocalised with eQTLS for genes enriched for expression in regions of the brain including the amygdala, cerebellum and cortex (Table A16).

Furthermore, we performed gene-set and tissue enrichment analyses using DEPICT [43, 44]. We found an association between the genetic variants affecting hypothalamus volume in females and the cardiovascular system (*P* = 5.54 *×* 10*^−^*^4^, Fig. A7, Tables A17, A18 & A19).

### Enrichment for gene expression in ovarian cell-types

To test for enrichment of gene expression at the ovarian cell type level, we used an in-house single-cell ovary atlas consisting of a combination of publicly available and in-house single-cell RNA-seq datasets (paper in preparation, Ferreira *et al.*, see Methods for further details). Using CELLECT [45], the in-house annotated single-cell RNA-seq count data was combined with our neuroendocrine IDP GWAS summary statistics to identify associated candidate aetiological cell types.

We identified statistically significant (*P <* 0.05 after BH FDR correction) enrichment of signal in each of the four neuroendocrine IDPs in gene regions specifically expressed in five ovarian cell types (Fig. 4A, Table A20). For example, the pituitary gland associated genetic variants discovered in the sex-combined sample were enriched for expression in genes specific to the early theca cells, theca interna, theca externa and the cumulus-oocyte complex. Variants associated with hypothalamus, hypothalamic grey matter and olfactory bulb volume, all in the sex-combined sample, and with hypothalamus volume in the female only sample, were enriched for expression in theca externa cell genes. Additionally variants associated with hypothalamic grey matter and olfactory bulb volume in the sex-combined sample were enriched for early theca cell expressed genes. Theca cells are key to the endocrine function of the ovaries, responding to LH released by the pituitary gland to generate testosterone and precursors of oestradiol [46]. We also found significant enrichment of genes expressed in the surface epithelium for genetic variants associated with hypothalamic grey matter and olfactory bulb volume in the sex-combined sample, and hypothalamus volume in the sex-combined and female only sample. The ovarian surface epithelium has both FSH and LH receptors and gonadotropins have been shown to stimulate ovarian surface epithelium cell proliferation across several species. Additionally, GnRH acts as an autocrine growth inhibitor for the ovarian surface epithelium [47].

**Fig. 4.**
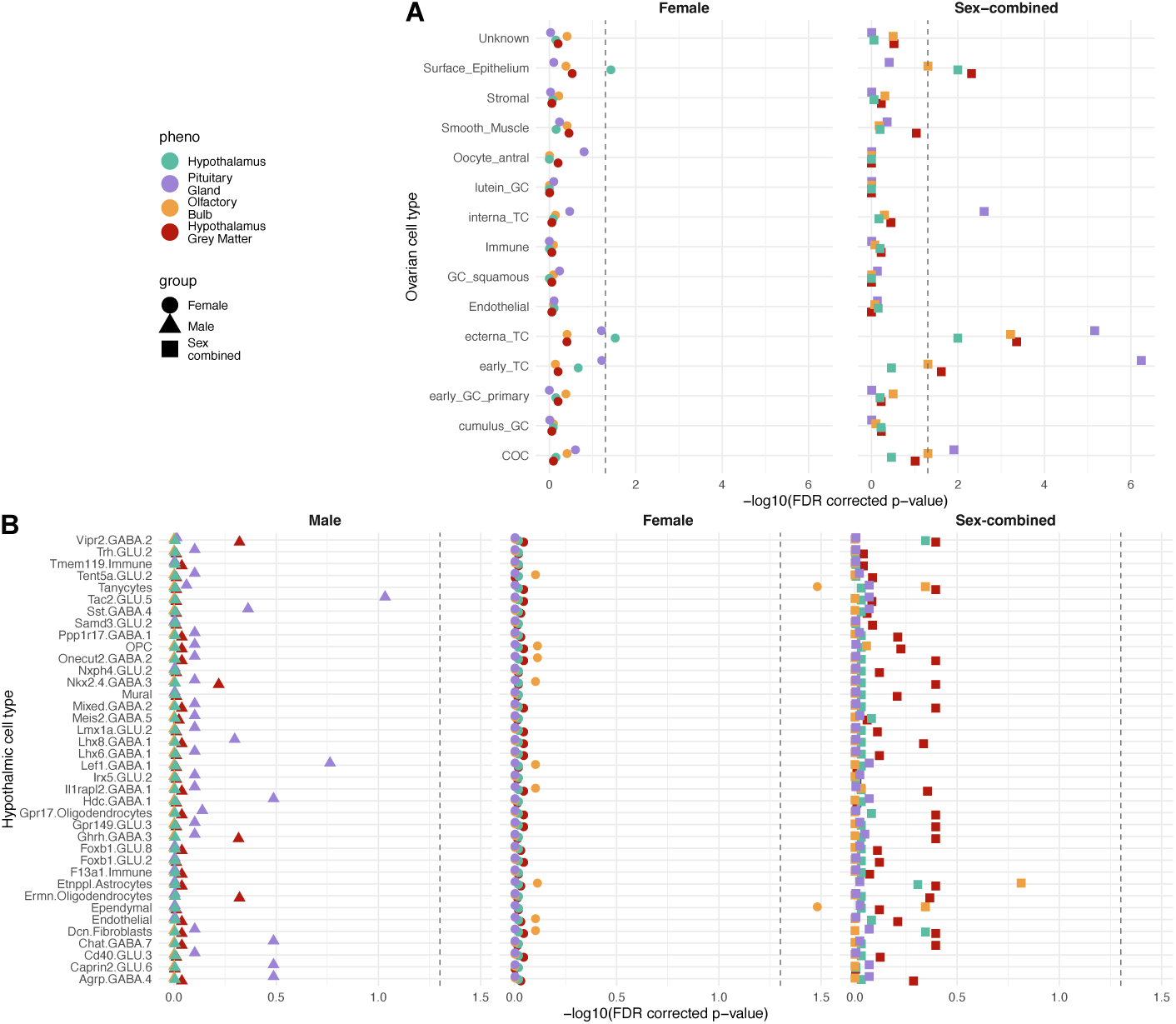
Enrichment for gene expression in ovarian and hypothalamic cell-types. Forest plots of CELLECT-based BH FDR corrected *P* -values signifying the genomic co-localisation between the discovered genetic loci and genes specifically expressed across ovarian (A) and hypothalamic (B) cells. The results are presented for analyses performed with the genetic variants with each of the four neuroendocrine IDPs across three popula-tion samples: volume of the hypothalamus (turquoise), pituitary gland (purple), mean of left and right olfactory bulbs (gold), and hypothalamic grey matter (red) across a female (circle), male (triangle) and sex-combined (square) sample. Significance thresholds (*P <* 0.05 after BH FDR correction) are indicated with a dashed grey line. A) Gene expression data from 15 ovarian cell types from five harmonised publicly available and four in-house expression datasets; GC, granulosa cell; TC, theca cell; COC, cumulus-oocyte complex. B) Gene expression data from 38 cell populations derived from a published human postmortem hypothalamus dataset [48].

### Enrichment for gene expression in hypothalamic cell-types

Using CELLECT [45] applied to published single-nucleus RNA-sequencing data from adult hypothalamus [48], we tested whether the genomic location of genes specifically expressed in any of 38 hypothalamic cell populations were enriched for association signals for any of the neuroendocrine IDPs (Table A21). Stratifying analysis based on variants identified in female, male and sex-combined GWASs, we identified two cell types exhibiting significant enrich-ment (*P <* 0.05 after BH FDR correction) for olfactory bulb volume in the female sample GWAS signal, namely tanycytes and ependymal cells (Fig.4B), two related cell types residing along the ventricles in the mediobasal hypothalamus. Tanycytes are a subtype of ependymal cells found in the third ventricle that project into the hypothalamus. There is evidence that tanycytes are involved in the release of GnRH due to the overlap in spatial distribution of the tanycytes and GnRH-releasing neurons [49]. Studies have shown that removal of tanycytes in rats impairs the release pulse of GnRH into the portal blood, and generation of the LH peak required for ovulation [50].

### Voxel-level genetic associations

We ran linear models at voxel-level to explore the effect of each of the genetic variants discovered during the four sex-combined GWAS in the rest of the brain (i.e., outside of the original neuroendocrine structures). The Jacobian determinant maps estimated from the nonlinear warp, aligning each individual to the standard template brain, were used as a measure of relative volume differences (expansion or contraction) at each voxel. The allele dosage for each of the 66 identified loci was regressed onto the Jacobian determinant maps, with the linear models including the same confound regressors used in the GWAS. Summary maps were calculated across the output t-statistic maps (one for each SNP) associated with each of the four IDPs, to represent for each voxel the proportion of discovered loci strongly associated (*t >* 3.5) with each volume. These models allowed for visualisation of the effect of genetic variation across the whole brain (Fig. 5, Fig. A8). The strongest effects on volume for each genetic variant appear to correspond with voxels within the neuroendocrine structures associated with that variant’s discovery. While this is not surprising, due to the circular nature of this analysis using these structures, this result provides reassuring validation of the method used.

**Fig. 5.**
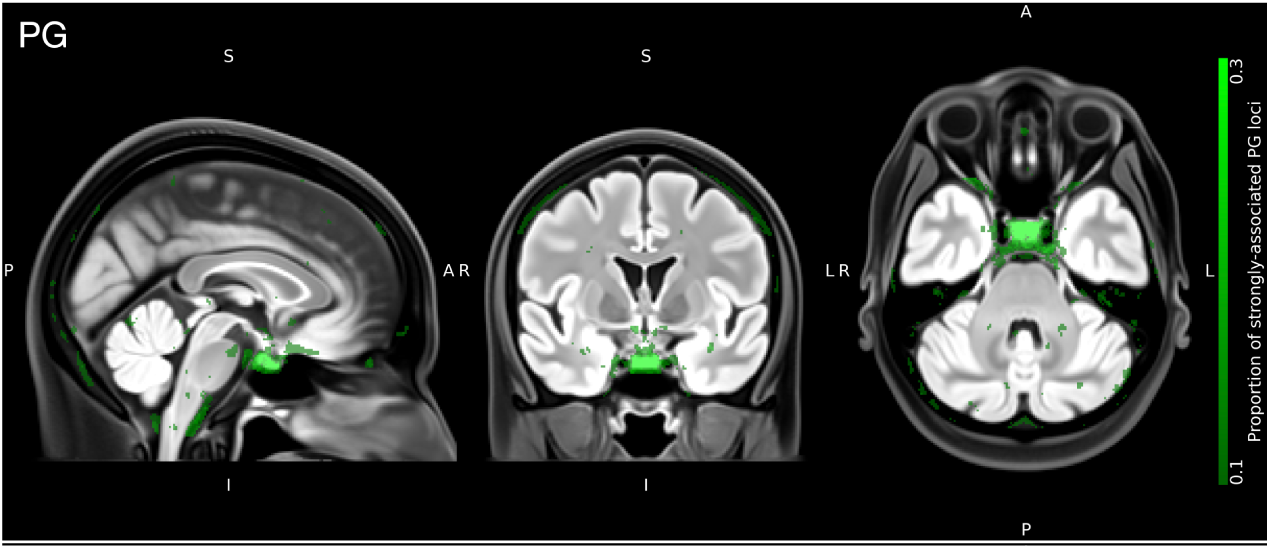
Voxel-wise associations of discovered genetic variants from pituitary gland volume GWAS. The summary map derived from the t-statistic maps overlaid on the OMM-1 T2-FLAIR reference template shows the proportion of the 16 variants identified in the GWAS of the pituitary gland volume in a sex-combined sample, that are also strongly-associated (*t >* 3.5) with the volume at a voxel location. The intensity range reflecting the strongly-associated proportion of SNPs in the summary map was set to 0.1 -0.3 (as shown in the colour bar) to highlight only voxels that are common in at least 10% of the identified PG SNPs.

Further, for each of the sets of SNPs associated with the four neu-roendocrine IDPs, the visualisation indicates genetic effect on brain regions additional to those already found via the IDPs. For example, the summary map of the pituitary gland volume associated loci (Fig. 5) shows associations with the cavernous and petrous sinuses and the basal vein of Rosenthal. These structures drain venous blood and thereby hormones from the pituitary gland. Associations can also be seen with the medial part of the H-shaped orbital sulcus, the hypothalamus, the amygdala and entorhinal cortex, as well as lobule IX of the cerebellum. Associations in the nucleus accumbens (colliculus nuclei caudati) and the midbrain structures can be seen in the hypothalamus and hypothalamic grey matter maps (Fig. A8). Surprisingly, large parts of the white matter, the fourth ventricle, and lateral ventricles around the foramen of Monro are associated too. Similar to the regions highlighted in the voxel-wise association of pituitary gland volume associated SNPs, we observe associations with venous structures, which drain (among other areas) circum-ventricular organs that lack the normal blood-brain barrier. Similarly to our finding of associations between SNPs from the neuroendocrine IDPs GWAS and additional brain structure phenotypes within the Big40 dataset, it is perhaps unsurprising to observe associations across structures of the brain given shared developmental and cellular compositions. The associations to the additional specified anatomical areas might rather point to areas of functionally related pathways.

## Methods

### Data

We used genetic information and multimodal brain imaging data from UKB [51]. UKB is one of the largest prospective epidemiological studies to date, containing genetic information as well as lifestyle and health measures from a cohort of *∼*500,000 individuals. Amongst other organ imaging efforts, UKB is acquiring brain MRI from 100,000 participants, with collection ongoing.

Here, we used MRI data from 37,330 UKB individuals (45-84 year age range) with usable T1-weighted (T1), T2-weighted fluid-attenuated inversion recovery (T2-FLAIR) and diffusion-weighted MRI (dMRI). T1 is one of the most commonly used sequences due to its strong contrast between grey matter, white matter, and cerebrospinal fluid, allowing a high-resolution depiction of basic brain anatomy. T2-FLAIR as a second structural modality provides strong contrast in some subcortical grey matter structures and the olfactory bulbs, as well as between white matter hyperintensities and normal-appearing white matter. dMRI provides information about local microstructural tissue properties and allows the estimation of a diffusion tensor, which adds information about axonal organisation. Scans had been acquired at one of three sites using identical 3T Siemens Skyra scanners and a standard 32-channel receive head coil. A detailed description of the acquisition protocol can be found in [51].

The complementary information provided by these spatially corresponding imaging modalities helps localisation of anatomical regions of interest (ROIs) and interpretation of results, and improves the accuracy when spatially normalising individuals to a template. Here, we used the recently released Oxford-MultiModal-1 (OMM-1) template [52] as the common space for alignment. The OMM-1 is a fully-unbiased and internally-consistent multimodal template that represents the average brain anatomy and appearance of 240 UKB subjects (randomly and uniformly sampled from the 50-55 year age group, 50% female). It comprises anatomically-corresponding volumes from the same modalities (T1, T2-FLAIR and DTI), which were also used to inform the construction process. Consequently, the OMM-1 provides optimal compatibility with respect to brain shape and appearance, and consistency for the current study population and task.

UKB has approval from the North West Multi-centre Research Ethics Com-mittee (http://www.ukbiobank.ac.uk/ethics/), which also covers the work in this study.

### Data preprocessing

The UKB brain imaging data had undergone extensive manual and automated quality control and preprocessing steps as described in [53]. In brief, the pipeline includes manual and semi-automated quality checks, gradient-distortion correction, cropping, brain extraction (removal of non-brain tissue such as the skull, scalp, and eyes), and intensity inhomogeneity correction. For each individual, the T2FLAIR and dMRI modalities are rigidly co-registered to the the space of the corresponding T1 image with FSL FLIRT [54]. Partial volume tissue segmentations are estimated for each of three tissue types (grey matter, white matter, and cerebrospinal fluid) with FSL FAST [55]. Tensors are fit to the dMRI data with FSL’s DTIFIT tool [56] after correcting for susceptibility and eddy current-induced distortions, and head motion, using FSL’s topup [57] and eddy [58, 59] tools.

### Image-derived phenotype extraction

We extracted Image-derived phenotypes (IDPs), i.e., volume measurements for our ROIs, in individuals’ native spaces through label propagation. This approach involved creating masks by segmenting anatomical ROIs in a common reference template, spatial normalisation (estimation of transformations) of all individuals to this reference template, and application of the inverse transformations to these masks to propagate them from the reference space to the original spaces of the individuals.

We first created initial binary masks for the three primary anatomical ROIs (pituitary gland, hypothalamus and olfactory bulbs) in the space of the OMM-1. We manually delineated the olfactory bulbs (left and right separately) and pituitary gland in the T2-FLAIR and T1 volumes of the OMM-1 population average template, respectively. These binary masks were modulated by the corresponding image intensities to account for partial volume effects, which provided spatial segmentation maps with values in the 0-1 range. Note that, in contrast to the posterior pituitary gland (neurohypophysis, formerly called ‘pars cerebralis’ of the pituitary), the anterior pituitary gland (adenohypophysis) and the pars intermedia (both formerly called ‘pars pharyngealis’ of the pituitary) are phylo-and ontogenetically not part of the brain but derivatives of Rathke’s pouch. Thus, the anterior pituitary is essentially an epithelial gland. The discrimination between posterior and anterior pituitary (including the pars intermedia) was not possible based on the available MRI dataset and would require more specific acquisition sequences. This has to be taken into consideration when interpreting the results.

A hypothalamus mask was obtained by transforming two existing atlases to OMM-1 space and combining them. The first hypothalamic atlas used here is the one constructed by Neudorfer *et al.* [60]. For the construction, two raters performed manual delineation in a high-resolution (initial resolution of 0.7mm upsampled to 0.25mm isotropic), high-contrast template constructed from 990 individuals in the Human Connectome Project [61] after registering and transforming them to MNI space. We resampled this binary hypothalamic mask into OMM-1 space using the transformations estimated during affine and nonlinear registrations between the MNI template and the T1 volume of the OMM-1 template. The second hypothalamus mask used was created by Pauli *et al.* [62]. They performed manual delineation in each of eight validation templates before combining them into a probabilistic segmentation map. Each of the validation templates was constructed from 81 individuals sampled from the same random set of 168 HCP individuals [61]. We resampled this probabilistic hypothalamic map into OMM-1 space using the transformations estimated during affine and nonlinear registrations of their study-specific group template to the T1 volume of the OMM-1 template. Both transformed atlases were merged in OMM-1 space by masking the probabilistic mask with the binary mask.

Next, we spatially normalised all individual brains to the reference space of the OMM-1 template. The brain-extracted T1 image of each individual was affinely (12 degrees of freedom) registered to the T1 volume of the OMM-1 with FLIRT [54]. This affine transformation was used to initialise the nonlinear registration with FSL’s MultiModal Registration Framework (MMORF) [63]. MMORF estimates one deformation field for all modalities and provides, from this, a derived Jacobian determinant map between an individual brain and the reference template. The deformation field provides a spatial mapping between anatomically corresponding ROIs and the Jacobian map provides a measure of expansion/contraction for each voxel. All three modalities, T1, T2-FLAIR and DTI, from both individuals and the template were used to drive the registrations for optimal registration accuracy. Hypothalamus, pituitary gland, and olfactory bulb masks were transformed from OMM-1 space to each individual’s native space by applying the corresponding inverse transformations with trilinear interpolation.

Finally, due to the intensity modulation and effects of interpolation, the transformed masks in subject space had values in the 0-1 range, where a mask value of 0 indicates that the voxel definitely is not part of the anatomical structure and a value of 1 indicates that the voxel definitely is part of the structure. Values in-between indicate partial volumes, e.g., a value of 0.5 indicates that half of the voxel contains the structure of interest. The transformed masks were thresholded at 0.3 and binarised as this provided a good trade-off between over-and undersegmentation (assessed visually). Using these binarised masks we calculated the volume (mm^3^) for each structure, producing three IDPs (pituitary gland volume, hypothalamus volume and combined olfactory bulbs volume). In addition, we extracted the grey matter (GM) volume of the hypothalamus from the estimated GM partial volume maps, producing our fourth IDP (hypothalamic grey matter volume).

### Image derived phenotype characterisation

The population was filtered using genotype principal component analysis to select for a population that represents the most densely populated well-mixed population, which here largely aligns with those who self-identify as White-European. To do so, in brief, we trained a random forest classifier to assign sample population labels using the 1000 Genomes [64] super-population labels (AFR=Africans, AMR=Admixed Americans, EAS=East Asians, Europeans=EUR, and South Asians=SAS) and genotype projected into principal component space. The resulting dataset contained 34,834 individuals. The mean and standard deviation of each IDP was calculated within this sample and in groups stratified by genetic sex, and age in bins of 10 years.

### Genome-wide association studies

We performed GWASs using an additive linear model implemented using REGENIE [65], a high-speed methodology that accounts for relatedness within the sample. The analysis was done in both sex-combined and sex-stratified samples. We regressed confound covariates from each phenotype using a generalised additive model in accordance with previous analysis: scan date and scan time, both with spline transformation applied; age; sex; head size; MRI table position in the *z* direction; brain centre of gravity in *x*, *y* and *z* dimensions; mean head motion. Categorical covariates were numerically encoded and all covariates were mean-centred prior to regression. Residuals were then inverse-rank normalised. The first 21 genetic principal components (PCs) were also provided as covariates to REGENIE. Resultant summary statistics files were filtered to remove genetic variants with a minor allele frequency *<*0.01, an imputation information score *<*0.3, a Hardy-Weinberg equilibrium *P* -value of *<*1 *×* 10^-15^ and any multi-allelic variants. We applied a genome-wide significance threshold of 5 *×* 10*^−^*^8^. We performed LDSC [10] to check for uncontrolled stratification among the resultant test statistics and to estimate SNP-based heritability. Considering the general regression models applied within REGENIE that account for relatedness, this heritability can be considered a lower-bound estimate. We identified significantly associated independent loci that are more than 10Mb apart using approximate conditional analysis implemented in GCTA-COJO [11]. Loci were manually annotated with the nearest coding gene and prior associations using ENSEMBL [66] and Open Targets Genetics [67].

### Sexual dimorphism

For each phenotype, we tested each significant (*P <* 5 *×* 10*^−^*^8^) lead variant identified across the different strata (sex-combined; and male-and female-specific;) for a sex-dimorphic effect by estimating the *t*-statistic:

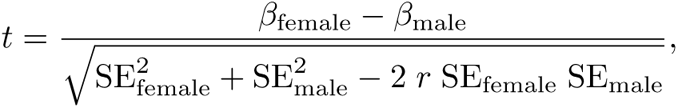

where *β*_sex_ and SE_sex_ (sex *∈ {*female, male*}*) are sex-specific effect sizes and SEs, respectively, and *r* is the genome-wide Spearman rank correlation between SNP effects in females and males. The *t*-statistic was estimated and the resulting *P* -value calculated [68] as implemented in the EasyStrata software [69].

### Exome analysis

#### Sequencing Quality Control

To evaluate an initial set of high quality variants we ran sample and variant level pre-filtering before calculating sample-level QC metrics. Following Karzcewski *et al.* [18], we then removed sample outliers using median absolute deviation (MAD) thresholds (*>* 4 MADs from the median). Finally, we applied a genotype-level filter using genotype quality (GQ *≥* 20), depth (DP *≥* 10), and heterozygote allele balance (AB *>* 0.2). After these filtering steps, the high-quality European call set consisted of 402,375 samples and 25,229,669 variants.

#### Variant Annotation

To annotate variants we used Variant Effect Predictor (VEP) v105, GENCODE v39 [70] with the LOFTEE v1.04 GRCh38 [71] and dbNSFP [72] plug-ins, annotating variants with CADD v1.6 [73], and REVEL [74] scores, and loss of function (LoF) confidence using LOFTEE. Full instructions to reproduce the annotation pipeline are provided within the VEP 150 LOFTEE repository (https://github.com/BRaVa-#genetics/vep105loftee). We then ran SpliceAI v1.3 [75] with the GENCODE v39 annotation file to ensure alignment between VEP and SpliceAI transcript annotations. For variant-specific annotations we defined ‘canonical’ transcripts as follows: set MANE Select [76] as the canonical where available, otherwise set canonical and restrict to protein-coding genes:

1. High confidence predicted LoF (pLoF): high-confidence LoF variants, as defined by LOFTEE (LOFTEE HC).
2. Damaging missense/protein-altering, at least one of:
  - Variant annotated as missense/start-loss/stop-loss/in-frame indel and (REVEL *≥* 0.773 or CADD *≥* 28.1 (or both)).
  - Any variant with SpliceAI delta score (DS) *≥* 0.2 where SpliceAI DS is the maximum of the set *{*DS AG, DS AL, DS DG, DS DL*}* for each annotated variant (where DS AG, DS AL, DS DG, and DS DL are delta score (acceptor gain), delta score (acceptor loss), delta score (donor gain), and delta score (donor loss), respectively).
3. Low-confidence LoF variants, as defined by LOFTEE (LOFTEE LC)
4. Other missense/protein-altering:
  - Missense/start-loss/stop-loss/in-frame indel not categorised in (2) (Dam-aging missense/protein-altering).
5. Synonymous: synonymous variants with SpliceAI DS *<* 0.2 in the gene (our ‘control’ set).

We selected the REVEL and CADD score thresholds based upon the American College of Medical Genetics and Genomics and the Association for Molecular Pathology’s (ACMG/AMP) PP3-moderate evidence criteria [77].

#### Rare variant association testing

Rare variant association testing was conducted using Scalable and Accurate Implementation of GEneralized mixed model (SAIGE, 1.1.9) [78], a mixed model framework that uses the saddlepoint approximation to calibrate the distribution of score test statistics in the presence of sample relatedness and case-control imbalance.

Beginning with our European-ancestry quality controlled subset of UKB we calculated a genetic relatedness matrix (GRM) using genotyping array data. We then LD pruned variants using the PLINK option --indep-pairwise 50 5 0.05 [79] and subset to 5,000 randomly selected markers to create a sparse GRM using SAIGE’s createSparseGRM.R.

For each trait we ran SAIGE step 1 to fit a null model with no genetic contribution using the sparse GRM with the first 20 genetic PCs and sex used as covariates. All default parameters were used except for --relatednessCutoff 0.05, --useSparseGRMtoFitNULL TRUE and --isCateVarianceRatio TRUE.

Finally, after fitting the null model we ran variant and gene-burden testing with SAIGE (step 2). All default parameters were used except for --maxMAF_in_groupTest=0.0001,0.001,0.01, --is_Firth_beta TRUE, --pCutoffforFirth=0.1 and --is_fastTest TRUE. For gene burden testing we used the annotations as previously defined, testing the following anno-tations groups: High confidence pLoF; Damaging missense/protein-altering; Other missense/protein-altering; Synonymous; High confidence pLoF or Dam-aging missense/protein-altering; and High confidence pLoF or Damaging missense/protein-altering or Other missense/protein-altering or Synonymous. The annotation results were also combined using a Cauchy combina-tion test [13] combining the tests for “High confidence pLoF”, “Damag-ing missense/protein-altering” and “High confidence pLoF AND Damaging missense/protein-altering” across all MAF limits. A conservative significance threshold of *P <* 6.25 *×* 10*^−^*^7^ was chosen accounting for multiple testing in number of genes and phenotypes considered.

### Phenome-wide association studies

We carried out PheWAS using ICD-10 codes from UKB, converted to phecodes as implemented in the PheWAS package in R [21]. Mean-centred age and sex and the first 21 genotype PCs were included as covariates and a significance threshold of P *<*0.05 following BH FDR correction was used. Further associa-tions were explored using the Big40 Oxford brain imaging genetics server. We extracted the marginal effect of the 66 variants significantly associated with the neuroendocrine IDPs on *∼*4,000 image derived phenotypes from multi-modal brain imaging [27]. The *P* -value significance threshold was adjusted for multiple testing using the Bonferroni method (*P <* 1.93 *×* 10*^−^*^7^).

### Shared genetics underlying neuroendocrine anatomical variation and reproductive traits

Here, we detail the analyses performed to investigate whether there is shared genetic effect on neuroendocrine IDPs, and reproductive traits. Reproductive traits examined included levels of testosterone, follicle stimulating hormone (FSH), luteinising hormone (LH), oestradiol and progesterone, alongside five phenotypes describing female infertility subtypes (all cause, anatomical cause, anovulatory, unknown cause excluding idiopathic infertility, and unknown cause including idiopathic infertility), and one male infertility phenotype. GWAS summary statistics of these traits were taken from Venkatesh *et al.* [39], in which further description of the phenotypes can be found. We used the summary statistics for the GWAS meta analysis of a European only sample. For all analyses apart from the enrichment for testosterone variants, we used the summary statistics from the meta analysis that excluded the UK Biobank data.

### Enrichment for testosterone variants

We used a one-sided fishers exact test, implemented in R, to calculate enrich-ment for testosterone variants (5) within our 66 neuroendocrine IDP associated variants. We obtained 254 lead variants associated with testosterone levels from the most-recent and largest meta-analysed GWAS of testosterone to our knowledge [39], among 1 million independent common variants across the genome [80].

### Genetic correlation

We estimated genetic correlations between the four neuroendocrine IDPs and hormone levels and infertility phenotypes using LDSC [10, 81]. Summary statistics were intersected with HapMap3 SNPs. The remaining set of variants were then used to estimate *r_g_*. Traits with a heritability Z-score *<*4 were excluded from *r_g_* estimation.

### Mendelian randomisation

Two-sample Mendelian randomisation was applied using the TwoSampleMR package in R [82, 83]. Bidirectional Mendelian randomisation was performed using the neuroendocrine IDP associated genetic variants as both exposure and outcome variables. Lead SNPs resulting from approximate conditional analysis applied in GCTA-COJO [11] (see methods above) were used as genetic instruments when using the neuroendocrine IDPs as the exposure variable. When using the reproductive traits as the exposure variable, genetic instruments were selected as the lead SNPs, defined using distance-based pruning as the variant with the lowest *P* -value within a 1Mb window of each genome-wide significant (*P <* 5 *×* 10*^−^*^8^) locus [39]. In both cases the internal clumping method was further applied using default parameters.

We applied five methods of meta-analyses: MR Egger, weighted median, inverse variance weighting, simple mode and weighted mode. Bonferroni multiple test correction was applied resulting in a corrected *P* -value significance threshold of 4.17 *×* 10^-3^. A secondary threshold was applied requiring the meta-analyses to include a minimum of five genetic instruments. Phenotype pairs were also tested for heterogeneity, horizontal pleiotropy, leave-one-out analysis and directionality [84–86].

### Colocalisation with reproductive phenotypes

Genomic position of the neuroendocrine IDP summary statistics was lifted from build hg19 to hg38 using UCSC’s liftOver [87]. Colocalisation was performed using the coloc package in R [88] applying a Bayesian approach under a single causal variant assumption. Genetic variants within a 100kb window around each lead variant associated with the neuroendocrine IDPs were used in the analysis. In line with recommendation of the coloc developers [89] we set the prior probabilities of genetic variants within a locus being associated with each trait trait to 1 *×*10*^−^*^4^, and to both traits to 1 *×*10*^−^*^6^. We assessed five hypotheses: *H*_0_ -no association with either trait; *H*_1_ -association with trait A but not trait B; *H*_2_ -association with trait B but not trait A; *H*_3_ -association with both traits but different causal variants; and *H*_4_ -association with both traits and the same causal variant. Associations were considered to be colocalised if 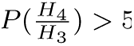 and *P* (*H*_4_) *>* 0.5.

### Colocalisation with tissue specific eQTLs

Colocalisation analysis with tissue specific eQTLs was performed using the coloc package in R [88] under a single causal variant assumption. The same priors and genomic window were used as above. Colocalisation was assessed with publicly available eQTL data from GTEx [32]. All genes with a transcriptional start site within a 1Mb region about the lead SNPs associated with the neuroendocrine IDPs were tested. Associations were considered to be colocalised if 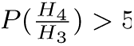 and *P* (*H*_4_) *>* 0.5 [90].

### Gene and Tissue Enrichment

To prioritise likely causal genes at the loci associated with our IDPs, and highlight enriched pathways and tissues where these genes are highly expressed, we performed gene-set and tissue enrichment analyses using DEPICT [43]. Briefly, DEPICT implements an integrative method that uses 14,461 gene sets reconstituted on the basis of expression data from 77,840 samples. Reconstituted gene sets that are enriched for genes in the associated loci are identified, with genes in different associated loci that have similar predicted functions prioritised. For tissue enrichment in DEPICT, microarray data from 37,427 human tissue samples are used to identify tissue and cell types in which genes from associated loci are highly expressed. A *P* -value threshold of 0.05 following BH FDR correction was used.

### Enrichment for gene expression in ovarian and hypothalamic cell-types

CELLEX (https://github.com/perslab/CELLEX)and CELLECT (https://github.com/perslab/CELLECT) were used to identify candidate aetiological cell types. In brief, CELLEX is used to compute robust cell-type expression specificity profiles by combining multiple expression specificity measures as described in Timshel *et al.* [45]. These estimates are used by CELLECT to test for association between genetic signal (heritability) taken from GWAS summary statistics, and cell-type expression specificity.

CELLEX was first applied to two datasets. Firstly gene expression measurements from 15 ovarian cell types from an in-house map comprising five harmonised publicly available ovarian gene expression datasets and four ovary samples processed in-house (paper in preparation, Ferreira *et al.*). Secondly from a dataset containing 38 hypothalamic cell populations derived from a human postmortem single-nucleus RNA-seq dataset [48], downloaded from https://storage.googleapis.com/linnarsson-lab-human/ adult human 20221007.loom.

CELLECT (version 1.1) was applied to GWAS summary statistics for each of the four neuroendocrine IDPs in sex-stratified and sex-combined analyses to find associations to the ovarian and hypothalamic cell-type expression specificity. MAGMA covariate analysis was used as the genetic prioritisation model [91]. CELLECT was run with default parameters, and significantly associated cell types were defined using a by-trait BH FDR corrected *P* -value threshold of *P <* 0.05.

### Voxel-level genetic associations

Similar to the approach taken in [6], we ran linear models to investigate the association between local variations in anatomical volume and allele dosage, of the 66 SNPs significantly associated with the neuroendocrine IDPs, at the voxel-level across the whole brain. The Jacobian determinant maps, which were obtained from the image registrations during the spatial normalisation processing step, were scaled by the determinant of the corresponding affine transformations to account for differences in brain size and used as a measure of volumetric variation. The intensity value of each voxel in a Jacobian determinant map indicates the ratio of the volume change in the registered subject at that voxel relative to the reference template, i.e., no change (=1), compression (*<*1) or expansion (*>*1). Allele dosages of SNPs that were previously identified as significant and the same covariates that were used during the GWAS were used as explanatory variables. Genetic dosage was extracted using qctools (https://www.chg.ox.ac.uk/~gav/qctool v2/).

The output t-statistic maps derived for each of the 66 SNPs were aggregated across their associated IDPs into four summary maps to highlight common brain regions strongly associated with an IDP. For example, for the PG we aggregated the t-statistic maps from 16 linear models, one for each variant identified as significantly associated with the PG IDP during the GWAS. These summary maps were created by taking the absolute values of the t-statistic maps followed by thresholding (*t >* 3.5), binarizing and averaging, to highlight strongly associated brain regions. Two expert neuroanatomists investigated these summary maps qualitatively. When interpreting the maps the potential statistical risks due to the circular nature of these analyses should be taken into consideration, i.e. no claims of significance were made.

## Discussion

We performed the first study to examine the genetic underpinnings of the anatomy of three neuroendocrine structures: the hypothalamus, pituitary gland and olfactory bulbs. Due to the functional nature of these structures within the hypothalamic-pituitary-gonadal axis, we hypothesised that the exploration of the genetics underlying the three neuroendocrine structures would lead to further understanding not only of their role in brain function, but also in broader endocrine and reproductive health. We extracted four quantitative image-derived phenotypes describing the volume of these largely unexplored regions across almost 35,000 individuals. In our analyses, we identified 66 independent loci significantly associated (*P <* 5 *×* 10*^−^*^8^) with one of these IDPs.

We found initial evidence of a shared genetic effect on both neuroendocrine anatomy and testosterone, one of the structures’ downstream products. There was significant enrichment (*P* -value = 9.89 *×* 10*^−^*^12^) of the genetic variants associated with the neuroendocrine IDPs for variants associated with testosterone. Five of the 66 lead genetic variants identified in the GWASs had a prior association to testosterone levels in or near to *REST*, *CENPW*, *CCND2*, *RNF2* and *PPM1A* [31]. In particular, the colocalisation of rs76895963 with an eQTL for *CCND2* across numerous tissues strongly suggests this is the the causal gene. *CCND2* has been robustly associated with testosterone across animal models and is known to have a role in fertility and reproductive processes [33–35]. Rare variant exome-wide analyses identified an association between hypothalamus volume and *STAB1*, a gene involved in iron homeostasis. Iron metabolism was also significantly associated with hypothalamic grey matter volume associated variants. Both iron metabolism and the related process of erythropoiesis are regulated by testosterone [24], with diseases affecting iron levels linked to reproductive dysfunction and infertility [23]. Taken together, these results provide initial evidence for an overlap in genetic effect between the volume of the four neuroendocrine regions and their function in production and regulation of testosterone. This is further supported by nominally significant genetic correlations between testosterone and pituitary gland, hypothalamus and olfactory bulb volumes.

We identified a possible link between genetic effect on neuroendocrine anatomy and the gonadal tissues in which their downstream products are active. We found enrichment of ovarian theca cells for genetic variants associated with pituitary gland volume. Theca cells, in response to LH produced by the pituitary gland as part of the hypothalamic-pituitary-gonadal axis, produce testosterone and precursors of oestradiol [46]. A genetic variant was also found downstream of *PPM1A*, a gene known to be involved in regulation of testosterone synthesis in Leydig cells in the testis [37]. In both cases this suggests a commonality to the genetics affecting hormone production across neuroanatomical and gonadal tissues. These two findings could represent shared pathways underlying the production and regulation of reproductive hormones, specifically testosterone, at different sites of the body. It could also be considered that the genetic links we are observing are caused by an effect of hormones produced in gonadal locations on the anatomy of the neuroendocrine structures. It would be ideal to analyse expression of genes across both gonadal and neuroanatomical tissues in the same individuals to aid the disentanglement of the causality underlying our observations, however no such datasets are currently available. Statistical analysis such as Mendelian randomisation could help further elucidate the relationship between the genetic variants effect on tissues that regulate hormones, and the hormone levels themselves, however we have thus far been underpowered to discover such causal relationships.

Given the androgen cascade controlled by the hypothalamic-pituitary-gonadal axis, there could still be underdiscovered links between anatomy of the neuroendocrine structures and additional reproductive hormones beyond testosterone. The pituitary gland is more directly involved in production of FSH and LH, yet we did not observe evidence for a genetic link between our phenotypes and the levels of these hormones beyond the ovarian cell enrich-ment. Currently, the discovery power of this analysis has been restricted by the limited sample size and resulting low SNP heritability of the hormone association studies. Particularly there is reduced statistical discovery power within the GWAS of FSH and LH, for which sample sizes in the GWAS which we used to look for genetic overlap were less than half that of testosterone. Furthermore, the genetic basis of testosterone has been more intensively studied than FSH and LH. Additional datasets with more accurate and comprehensive measurement of hormones in a healthy population are required to be able to further elucidate the link between neuroanatomical structures of the endocrine system and reproductive hormones. The addition of temporal collection of hormonal data would also allow an understanding of intrapersonal variation that could be affecting statistical power of genetic discovery, and hence discovery of genetic overlap between the phenotypes.

We identified a possible shared role of the genetic variants associated with neuroendocrine anatomy on not just production of reproductive hormones, but also on mechanistic function of their release. Olfactory bulb volume associated variants were enriched in two cell types across hypothalamic cells tested, the ependymal cells, and their subtype, the tanycytes. Notably tanycytes have previously been shown to influence the release of GnRH [49]. Further, employment of voxel-wise association studies identified association between the identified genetic variants and areas around the cavernous and petrous sinuses and the basal vein of Rosenthal, which are responsible for draining venous blood and hence hormones from the pituitary gland. This suggests a possible role of the discovered genetic variants not only in hormone production but also in the more mechanistic role of their release into the blood. The voxel-wise analysis allows us to identify regions of the brain affected by our genetic discovery set not captured by the quantitative image-derived phenotype summary measures that are more commonly used within genetics or clinical practice. It must be acknowledged that associations with the original neuroendocrine IDPs are inherently circular and one must be cautious when interpreting results, restricting only to structures assumed to be independent of the original phenotype measures. In this way it is still valuable as a method for visualising and exploring potential pleiotropy, and identifying regions of interest with potential for further investigation through the extraction of novel descriptive image-derived phenotypes for use in statistical genetics analyses.

The GWAS further identified several genetic variants with prior association to menstrual timing, particularly menarche and menopause. Particularly two genetic variants within the *STX6* gene were identified to have both had prior associations to the age at menopause. We also identified sicca syndrome to be associated with genetic variants associated with hypothalamic grey matter volume. Sicca syndrome is a condition that is largely diagnosed at onset of menopause and causes dryness in numerous glands [26]. It could be considered that the brain imaging that underlies the image-derived phenotypes was taken for many of the female participants at an age of substantial reproductive disruption -the perimenopausal and menopausal age. There is preliminary evidence that this process causes changes to neurological structures [92]. This could be leading us to discover some genetic effects that are specific to this time point. This could also account for some of the sexual dimorphism in genetic effects that we observe, which might be in part driven by the reproductive ageing process affecting the sexes differently. Work targeting the topic of sex-specific ageing more directly is needed in datasets that have additional phenotypic collection about menopause and associated hormonal changes. In parallel, further work using an imaging dataset of individuals around their reproductive peak could reveal more about the link between brain structure and reproductive function.

The extraordinary opportunity presented in the UKB dataset, to study the biology underlying common variation in anatomy through the examination of healthy population imaging coupled with genetics at a large scale, is unique. However, its uniqueness also presents a challenge in replication. The UKB is commonly acknowledged to represent a specific sociodemographic population, here particularly pertinent is the cohort’s older age. This might have affected the results as it is well documented that brain structure changes with advancing age. Ideally, replication would be conducted to confirm our results and assess the transferability of the results to cohorts of different population composition. However, there is currently a lack of accessible population-scale imaging datasets of non-disease specific individuals, especially those of ancestry outside of those that align with individuals who self-identify as “White European”. The creation of new datasets with equally rich phenotyping, but additional increased diversity, would allow for replication and expansion of our knowledge beyond a single population. Additional datasets would further enable the proposal of novel questions that expand our understanding of the connection between neuroanatomical and endocrine systems.

## Data Availability

Data used within the manuscript is available upon application to the UK Biobank.

## Appendix A

**Table A1** Mean volume (±standard deviation) of each neuroendocrine structure in the cohort stratified by age and sex.

**Fig. A1.**
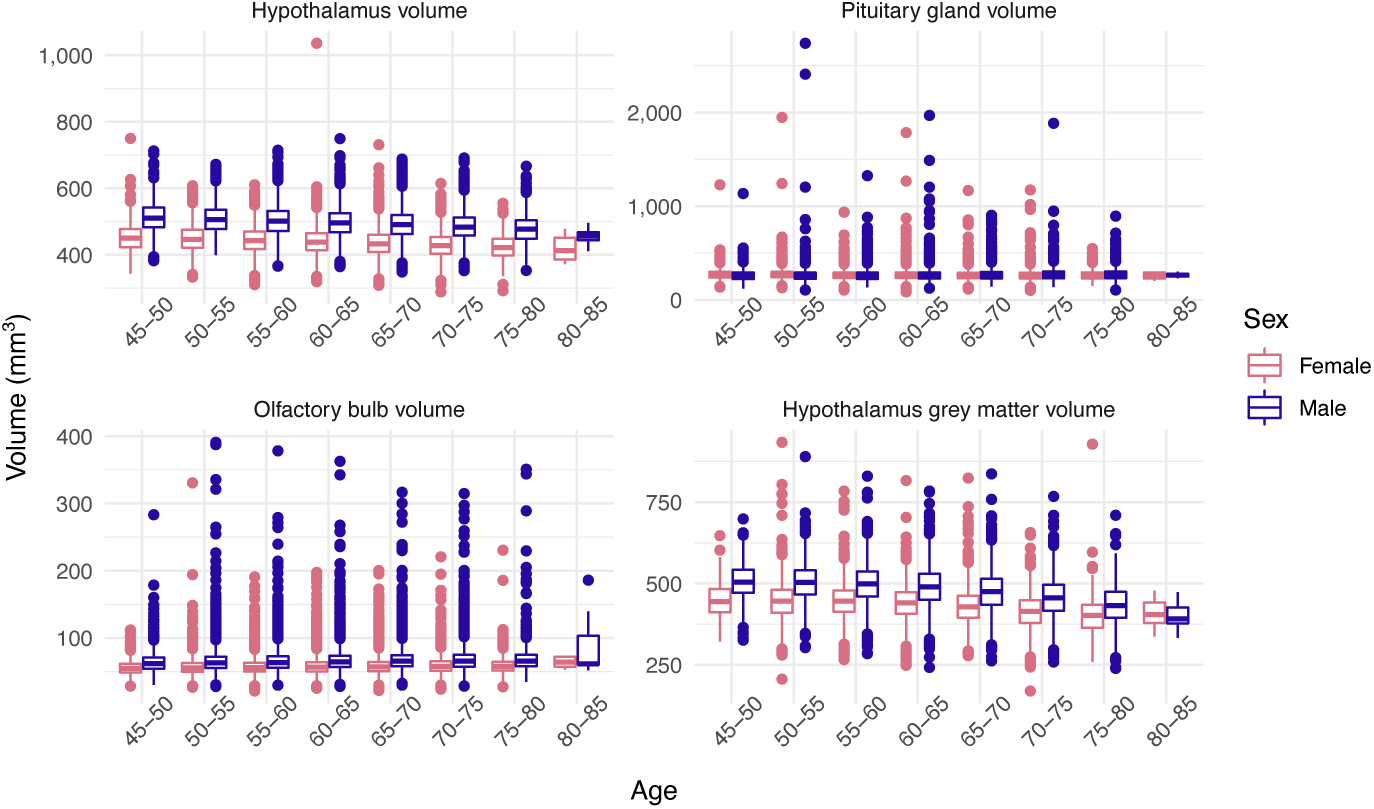
Distribution of neuroendocrine volumes across sex and age. The volumes (mm^3^) of the hypothalamus, pituitary gland, mean across left and right olfactory bulbs, and hypothalamic grey matter across age groups and sexes (as defined by genetic sex) within the study sample.

**Table A2** All statistically significant associations (*P <* 5 *×* 10*^−^*^8^) from the four genome-wide association studies of the volume of the hypothalamus (HT), hypothalamic grey matter (HT-GM), pituitary gland (PG) and olfactory bulbs (OB). For each the following data is presented: nearest coding gene assigned using OpenTargets Genetics; rsid; chromosome (CHROM); genomic position (GENPOS) in GRCh37 coordinates; reference allele (A0); effect allele (A1); effect allele frequency (A1FREQ); info score (INFO); sample size (N); Effect size; SE; and p-value.

**Table A3** All statistically significant associations (*P <* 5 *×* 10*^−^*^8^) from the four genome-wide association studies of the volume of the hypothalamus (HT), hypothalamic grey matter (HT-GM), pituitary gland (PG) and olfactory bulbs (OB). For each the following data is presented: nearest coding gene assigned using OpenTargets Genetics; rsid (ID); chromosome (CHR); phenotype; *P* -value; prior associations to reproductive traits; and prior associations to brain-related phenotypes, all assigned using OpenTargets Genetics [67] and ENSEMBL [66].

**Fig. A2.**
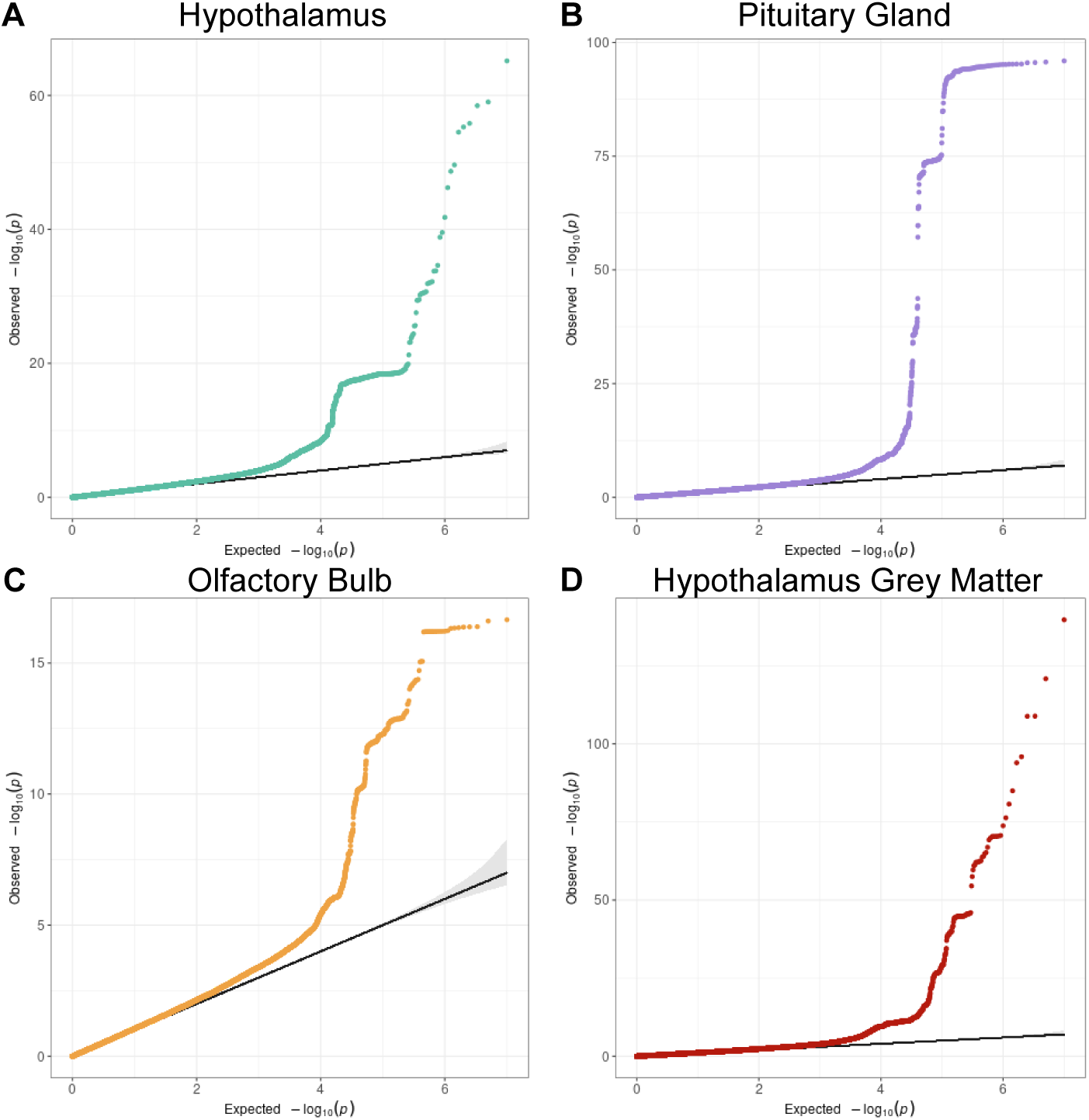
Quantile-quantile plots for GWAS of neuroendocrine IDPs. Plot of expected (x-axis) vs observed (y-axis) *−log*_10_(*P* -values) ranked for each SNP tested across the four genome wide association studies: A) hypothalamus; B) pituitary gland; C) olfactory bulb; and D) hypothalamic grey matter.

**Table A4** Linkage score disequilibrium metrics for the four GWAS of the neuroendocrine IDPs: the hypothalamus (HT), the pituitary gland (PG), the mean of the left and right olfactory bulbs (OB) and the hypothalamic grey matter (HTGM). Metrics include lambda genomic control (GC), linkage disequilibrium score (LDSC) intercept, LDSC intercept standard error (SE), LDSC ratio, LDSC ratio SE and LDSC intercept *P* -value.

**Fig. A3.**
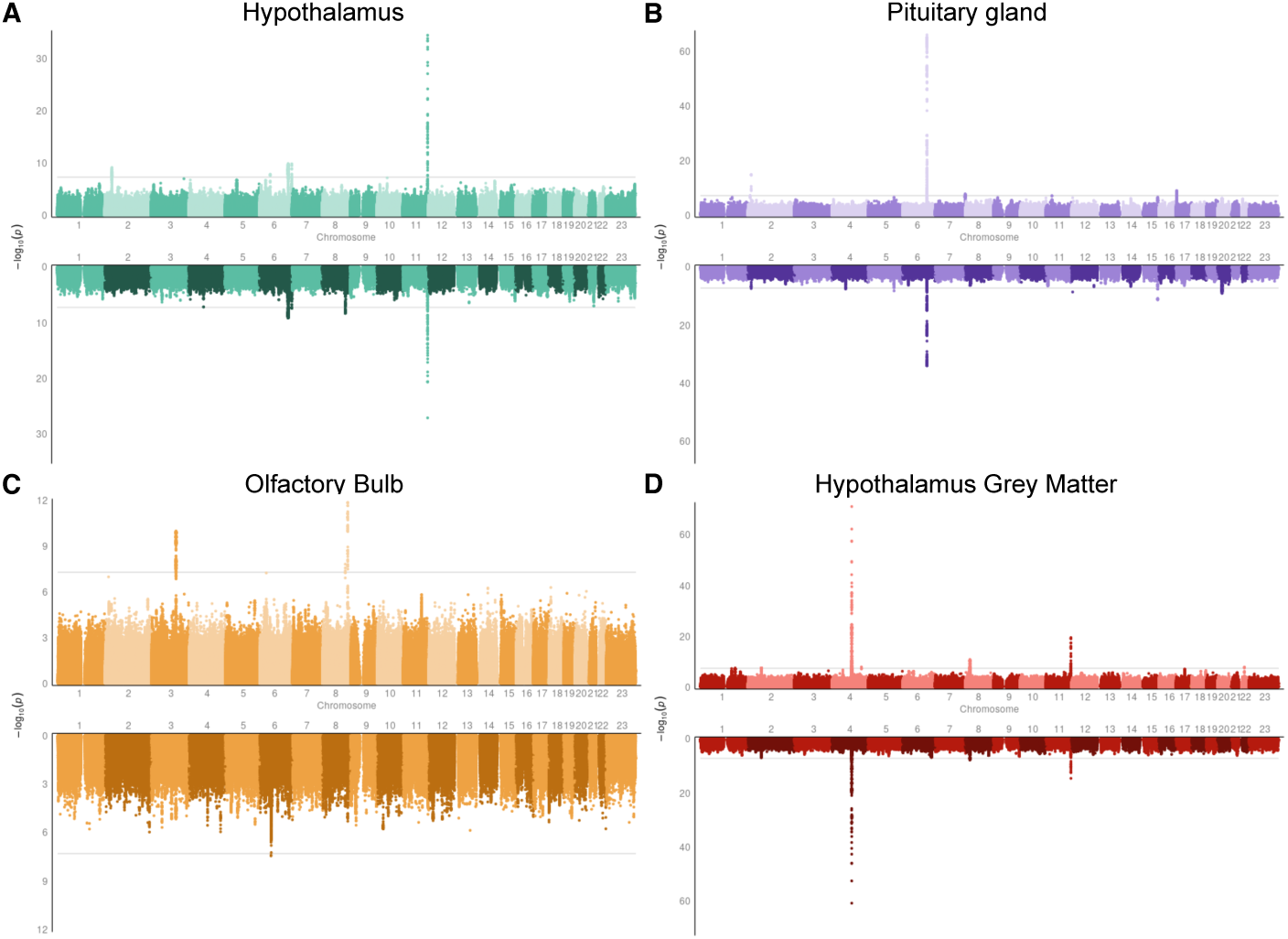
Genome-wide association studies of the four neuroendocrine IDPs in female and male samples. Miami plots presenting the results of genome-wide association studies of the volume of the hypothalamus (A), pituitary gland (B), olfactory bulb (C) and hypothalamic grey matter (D) in a female sample (n= 18,487, upper, light) and male sample (n= 16,347, lower, dark).

**Table A5** SNPs with statistically significant sexual dimorphism in genetic effect on the neuroendocrine IDPs. For each SNP the following information is listed: neuroendocrine IDP with which the genetic variant is associated, SNP rsid, chromosome (CHR), genomic position (POS), other allele (OA), effect allele (EA), effect allele frequency (EAF), effect size (BETA), SE, and *P* -value (P) in a male and female sample and the *P* -value from the sexual dimorphism test (*P*_DIFF_).

**Table A6** Results of phenome-wide association studies using genetic variants associated with hypothalamus volume and ICD10 codes from the UKB converted to phecodes. For all associations with phenotypes where there is a case sample size *>* 5, the following results are listed: rsid; phecode (phenotype), phecode description, phecode group, effect size (beta), SE, OR, *P* -value, BH FDR adjusted *P* -value (fdr adj p), case count, and control count.

**Table A7** Results of phenome-wide association studies using genetic variants associated with pituitary gland volume and ICD10 codes from the UKB converted to phecodes. For all associations with phenotypes where there is a case sample size *>* 5, the following results are listed: rsid; phecode (phenotype), phecode description, phecode group, effect size (beta), SE, OR, *P* -value, BH FDR adjusted *P* -value (fdr adj p), case count, and control count.

**Table A8** Results of phenome-wide association studies using genetic variants associated with olfactory bulb volume and ICD10 codes from the UKB converted to phecodes. For all associations with phenotypes where there is a case sample size *>* 5, the following results are listed: rsid; phecode (phenotype), phecode description, phecode group, effect size (beta), SE, OR, *P* -value, BH FDR adjusted *P* -value (fdr adj p), case count, and control count.

**Fig. A4.**
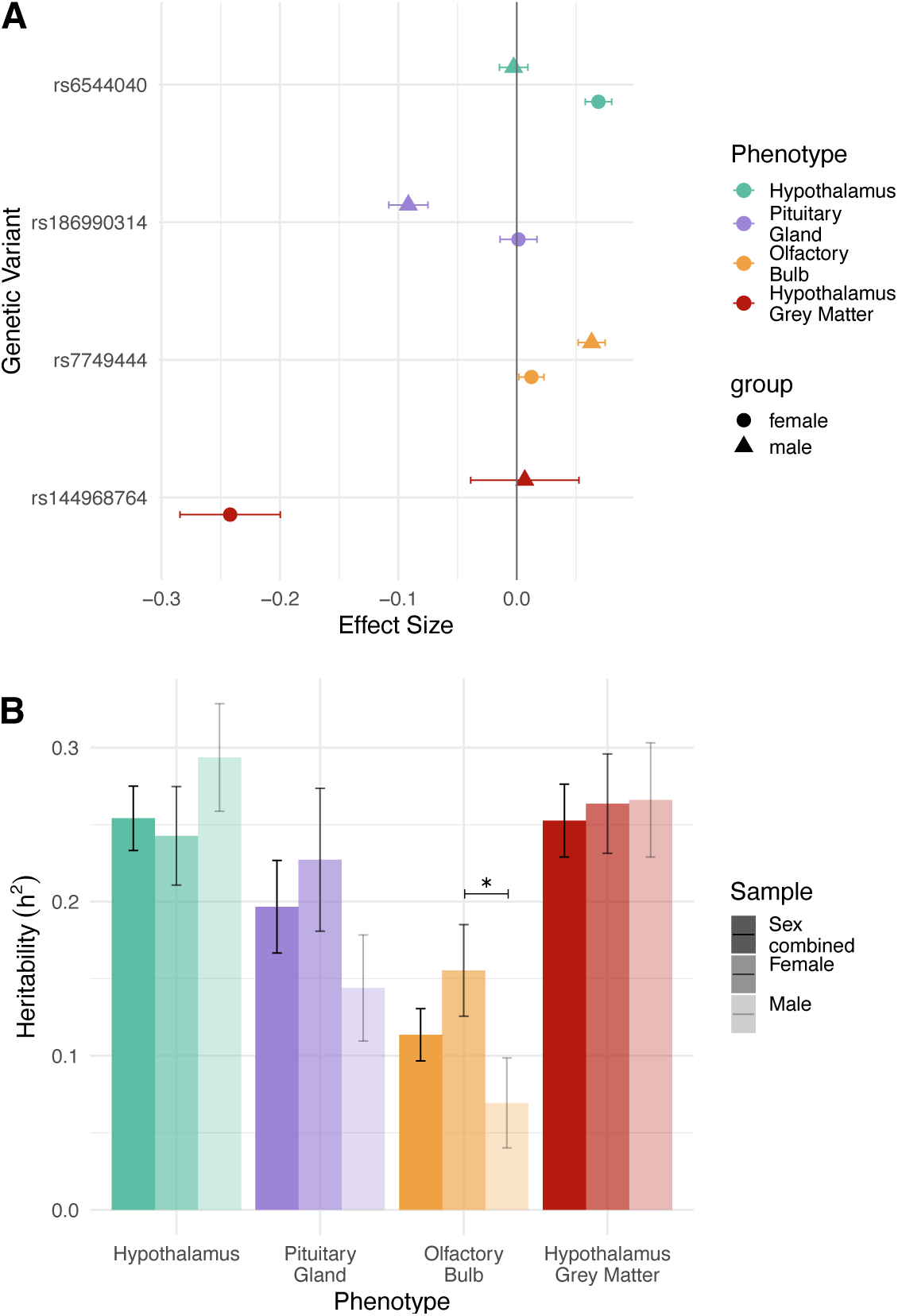
Sexual dimorphism in genetic effect on neuroendocrine IDPs. A) The genetic effect of SNPs with statistically significant sexual dimorphism in genetic effect tested across all SNPs significantly associated in the female, male or sex-combined samples across the GWAS of the four neuroendocrine IDPs: hypothalamus (turquoise), pituitary gland (purple), olfactory bulb (gold) and hypothalamic grey matter (red). Genetic effect on females is represented as a circle, genetic effect on males is represented by a triangle. SE is presented as a horizontal line about the point. B) The heritability (*h*_SNP_^2^) of the volume of the four neuroendocrine IDPs in the sex-combined (darkest), female (medium opacity) and male (lightest) samples as calculated in LDSC [10]. Statistically significant differences between males and females are highlighted with an asterisk.

**Fig. A5.**
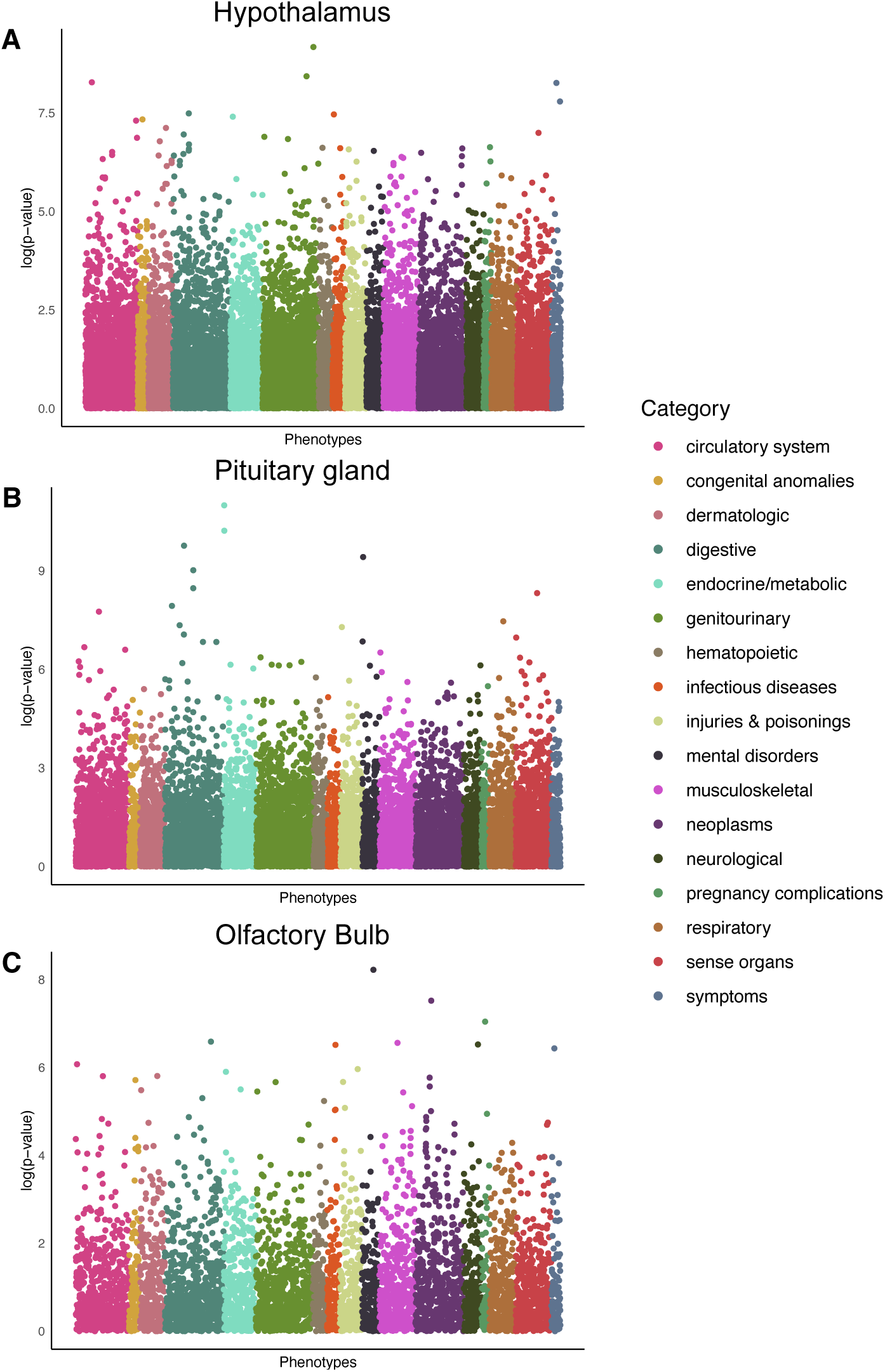
Phenome-wide association studies using broader clinical phenotypes. Manhattan plot presenting the *−* log_10_(*P* -values) from a PheWAS between the UKB ICD10-codes converted to phecodes and the genetic variants identified as associated with the volume of A) hypothalamus; B) pituitary gland; and C) olfactory bulbs. Phecodes are categorised into broader categories represented by different colours.

Table A9 Results of phenome-wide association studies using genetic variants associated with hypothalamic grey matter volume and and ICD10 codes from the UKB converted to phecodes. For all associations with phenotypes where there is a case sample size > 5, the following results are listed: rsid; phecode (phenotype), phecode description, phecode group, effect size (beta), SE, OR, P-value, BH FDR adjusted P-value (fdr adj p), case count, and control count.

**Fig. A6.**
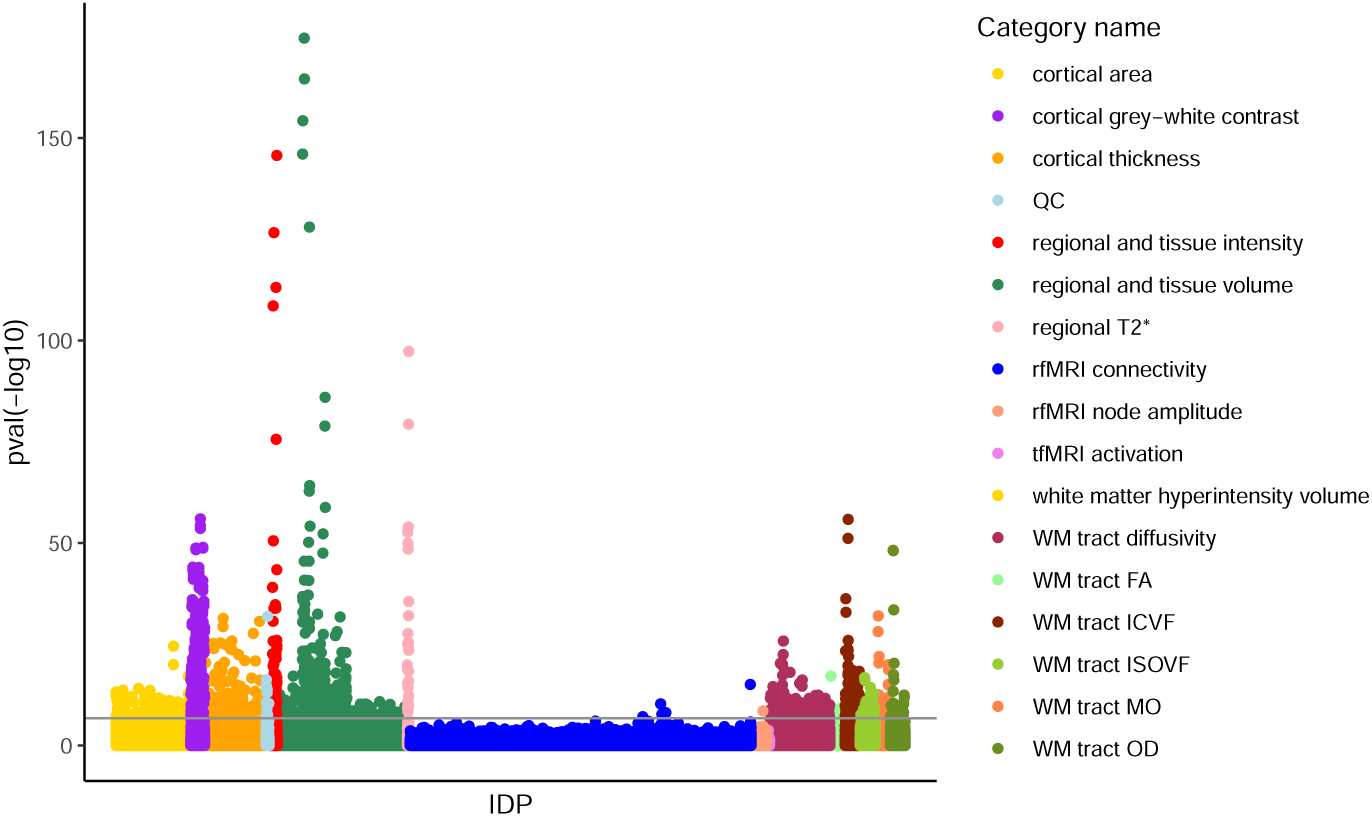
Phenome-wide association study of image-derived phenotypes from Big40 database. Manhattan plot presenting the *−log*_10_(*P* -values) from a phenome-wide association study utilising image-derived phenotypes from multi-modal magnetic resonance imaging within the UKB stored within Big40 -a database of genetic effect on *∼*4000 image-derived phenotypes from multimodal brain imaging [27], and the 66 genetic variants discovered across the four GWAS of the hypothalamus, pituitary gland, olfactory bulb and hypothalamic grey matter volume in a sex-combined sample.

**Table A10** All 1,140 statistically significant associations (*P <* 1.93 *×* 10*^−^*^7^) between the genetic variants associated with hypothalamus volume in a sex-combined sample and the Big40 database IDPs. The IDP ID (IDP), chromosome (chr), rsid, genomic position (pos), effect allele (a1), reference allele (a2), effect size, standard error (se), pvalue, IDP short name and IDP category name are listed.

**Table A11** All 82 statistically significant associations (*P <* 1.93 *×* 10*^−^*^7^) between the genetic variants associated with pituitary gland volume in a sex-combined sample and the Big40 database IDPs. The IDP ID (IDP), chromosome (chr), rsid, genomic position (pos), effect allele (a1), reference allele (a2), effect size, standard error (se), pvalue, IDP short name and IDP category name are listed.

**Table A12** All 22 statistically significant associations (*P <* 1.93 *×* 10*^−^*^7^) between the genetic variants associated with olfactory bulb volume in a sex-combined sample and the Big40 database IDPs. The IDP ID (IDP), chromosome (chr), rsid, genomic position (pos), effect allele (a1), reference allele (a2), effect size, standard error (se), pvalue, IDP short name and IDP category name are listed.

**Table A13** All 1,228 statistically significant associations (*P <* 1.93 *×* 10*^−^*^7^) between the genetic variants associated with hypothalamic grey matter volume in a sex-combined sample and the Big40 database IDPs. The IDP ID (IDP), chromosome (chr), rsid, genomic position (pos), effect allele (a1), reference allele (a2), effect size, standard error (se), pvalue, IDP short name and IDP category name are listed.

**Table A14** Genetic correlation between the four neuroendocrine IDPs and reproductive traits. The reproductive traits include testosterone (test), follicle stimulating hormone (FSH), luteinising hormone (LH), oestradiol (oest) and progesterone (prog) levels, female all cause infertility (infertility1), female anatomical cause infertility (infertility2), female anovulatory infertility (infertility3), female unknown cause infertility excluding idiopathic infertility (infertility4), female unknown cause infertility including idiopathic infertility (infertility5), and male infertility (male infert).

**Table A15** Results from Mendelian randomisation analyses of relationships between the volume of the hypothalamus, pituitary gland, olfactory bulb and hypothalamic grey matter volume, and reproductive phenotypes including: testosterone (test), follicle stimulating hormone (FSH), luteinising hormone (LH), oestradiol (oest) and progesterone (prog) levels, female all cause infertility (infertility1), female anatomical cause infertility (infertility2), female anovulatory infertility (infertility3), female unknown cause infertility excluding idiopathic infertility (infertility4), female unknown cause infertility including idiopathic infertility (infertility5), and male infertility (male infert).

**Table A16** Genetic variants significantly colocalised (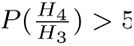 and *P* (*H*_4_ ) *>* 0.5) with tissue-specific eQTLs within the GTEx dataset [32].

**Fig. A7.**
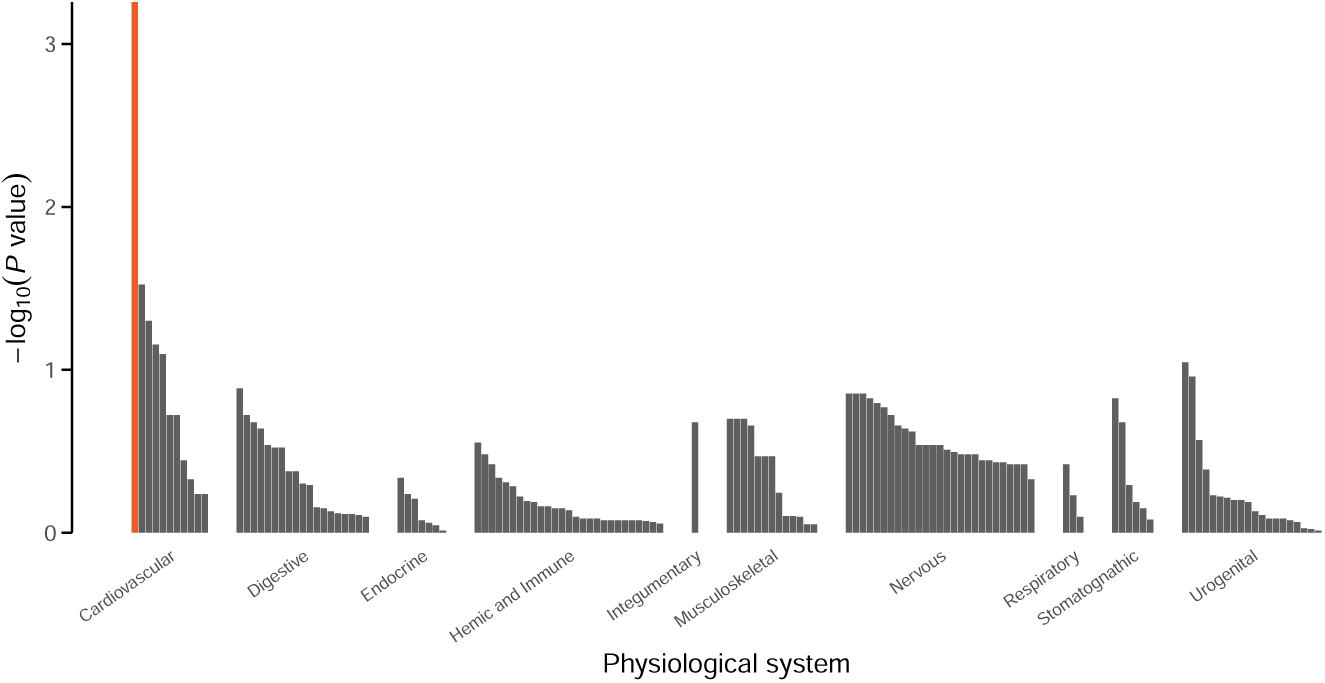
Pathway enrichment of hypothalamus associated genetic variants in a female sample. Significance of enrichment of the genetic variants associated with hypothalamus volume in females across different pathways within physiological systems, assessed using DEPICT [43]. Significant associations (*P <* 0.05 after BH FDR correction) are highlighted in red.

**Table A17** Gene prioritisation of genetic variants associated with the four neuroendocrine IDPs. Prioritisation was assessed using DEPICT [43] and BH FDR correction used with a significance threshold of 0.05.

**Table A18** Pathway enrichment of genetic variants associated with the four neuroendocrine IDPs. Enrichment was assessed using DEPICT [43] and BH FDR correction used with a significance threshold of 0.05.

**Table A19** Tissue enrichment of genetic variants associated with the four neuroendocrine IDPs. Enrichment was assessed using DEPICT [43] and BH FDR correction used with a significance threshold of 0.05.

**Table A20** All results from enrichment analysis for gene expression in ovarian cell-types using CELLEX and CELLECT [45]. Gene expression data represents 15 cell populations derived from five harmonised publicly available ovarian datasets and four ovary samples processed in-house; GC, granulosa cell; TC, theca cell; COC, cumulus-oocyte complex.

**Table A21** All results from enrichment analysis for gene expression in hypothalamic cell-types using CELLEX and CELLECT [45]. Gene expression data represents 38 cell populations derived from a published human postmortem hypothalamus dataset [48].

**Fig. A8.**
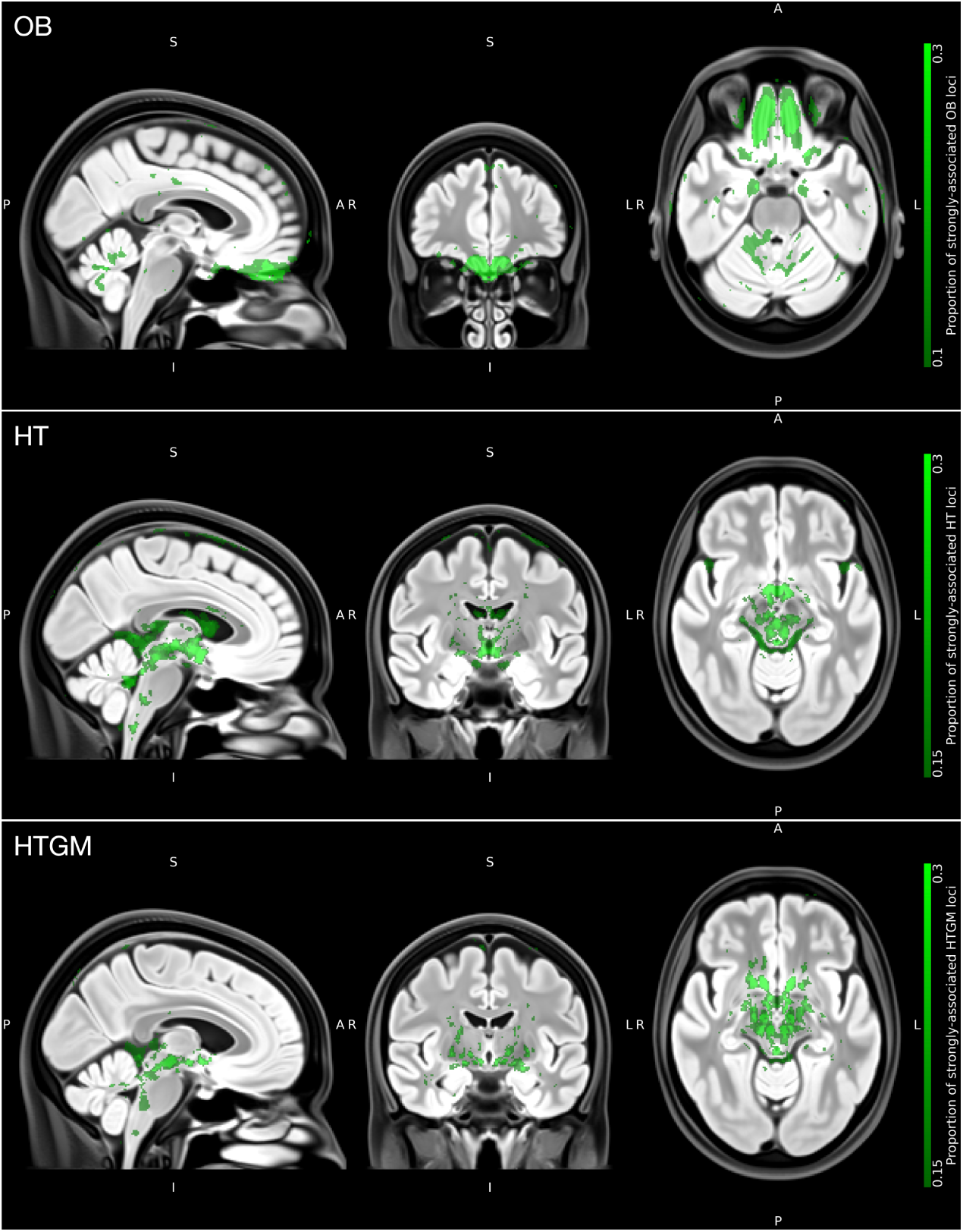
Voxel-wise associations of neuroendocrine IDP associated genetic variants. Summary maps overlaid on the OMM-1 reference template showing the proportion of strongly-associated loci for each voxel, from each of the three sets of loci identified in GWAS (olfactory bulbs (OB), hypothalamus (HT), and hypothalamic grey matter (HTGM) volume in a sex-combined sample).

